# The effect of vaccination on transmission of SARS-CoV-2 (COVID-19): a rapid review

**DOI:** 10.1101/2022.12.09.22283255

**Authors:** Jessica Williams, Sasha Barratte, Tom Winfield, Lauren Elston, Katie McDermott, David Jarrom, Elise Hasler, Caron Potter, Ruth Lewis, Alison Cooper, Adrian Edwards

## Abstract

This is an update (literature search up to 15 March 2022) of a <u>rapid review</u> examining whether vaccination against SARS-CoV-2 (COVID-19) affects transmission of SARS-CoV-2.

Streamlined systematic methodologies were used to accelerate the review process.

The update identified 17 additional studies: 6 studies reported on transmission and 11 studies reported viral load. There was high heterogeneity across studies, which varied in design, participant characteristics and SARS-CoV-2 variants reported. Evidence from this update supports previous findings that that transmission of Omicron and Delta variants is lowest in booster-vaccinated people, followed by fully vaccinated people, with the highest rate of transmission in unvaccinated people. Additionally, some studies compared transmission between different variants or sub-variants; risk of transmission appears to be higher with Omicron than Delta, regardless of vaccination status.

**Funding statement:** Health Technology Wales was funded for this work by the Wales COVID-19 Evidence Centre, itself funded by Health and Care Research Wales on behalf of Welsh Government.

## Wales COVID-19 Evidence Centre (WCEC) Rapid Review

#### Rapid Review Details

**Review conducted by:** Health Technology Wales

**Review Team:** Jessica Williams, Sasha Barrate, Lauren Elston, Katie McDermott, Tom Winfield, David Jarrom, Elise Hasler, Carron Potter

**Review submitted to the WCEC:** March 2022

**Rapid Review report issued by the WCEC:** November 2022

**WCEC Team:** Adrian Edwards, Alison Cooper, Ruth Lewis, Micaela Gal, Rebecca-Jane Law, Jane Greenwell and Ffion Coomber involved in drafting the Topline Summary, review and editing.

**This review should be cited as:** RR00054. Wales COVID-19 Evidence Centre. A rapid review of the effect vaccination on Sars-CoV-2 (COVID-19) transmission. March 2022 update.

**Disclaimer:** The views expressed in this publication are those of the authors, not necessarily Health and Care Research Wales.?The WCEC and authors of this work declare that they have no conflict of interest.

#### FULL REPORT

##### TOPLINE SUMMARY

###### What is a Rapid Review?

Our rapid reviews use a variation of the systematic review approach, abbreviating or omitting some components to generate the evidence to inform stakeholders promptly whilst maintaining attention to bias. They follow the methodological recommendations and minimum standards for conducting and reporting rapid reviews, including a structured protocol, systematic search, screening, data extraction, critical appraisal, and evidence synthesis to answer a specific question and identify key research gaps. They take 1- 2 months, depending on the breadth and complexity of the research topic/ question(s), extent of the evidence base, and type of analysis required for synthesis.

This review is closely linked to a prior Rapid Review (RR) conducted by Wales COVID-19. Evidence Centre (WCEC) and published as: What is the risk of SARS-CoV-2 transmission in vaccinated populations? Report number - RR00012 (November 2021)

###### Background / Aim of Rapid Review

The COVID-19 vaccination programme has been successful in reducing the impact of severe COVID-19 disease on hospitalisation, morbidity and mortality. However, the effectiveness of COVID-19 vaccines against transmission is less clear, in particular for people with milder or asymptomatic infection, or in an era of new variants. We previously conducted a RR that aimed to examine evidence on the transmission risk of SARS-CoV-2 from vaccinated people to unvaccinated or vaccinated people (Report number - RR00012, November 2021). At the time of this previous review, the UK Health Security Agency (UKHSA) were setting up a living rapid review on the effect of COVID-19 vaccination on transmission of SARS-CoV-2. This Living RR was continued until January 2022 when the last update search was conducted (UKHSA, 2022). This RR represents an update (search up to 15 March 2022) of the last version of the UKHSA living RR, and addresses the following review questions:

- Does vaccination against COVID-19 affect transmission of SARS-CoV-2 to others, in the subgroup of people who contract COVID-19 post-vaccination?
- How does risk of onward transmission vary with vaccine type, completion of the vaccination course, duration after vaccination, at different baseline community transmission levels and SARS-CoV-2 variant in the vaccinated person?

###### Extent of the evidence base

The update identified 17 additional studies: 6 studies reported on transmission and 11 studies reported viral load. One additional ongoing RCT was also identified (estimated publication date December 2024.)

###### Recency of the evidence base

The update review included evidence published between 12 January 2022 (end date of last search) and 15 March 2022.

###### Main findings

- The main conclusions from the previous review were not changed from the inclusion of 17 additional studies in this update.
- Fully and booster-vaccinated cases (infected persons) transmit SARS-CoV-2 less than unvaccinated cases, particularly for pre-Delta and Wild-type variants. This difference diminishes with time since vaccine dose, particularly for the Delta and Omicron variants.
- Studies of viral load showed similar findings, with most pre-Delta studies showing fully vaccinated cases had larger Ct values or lower viral loads than unvaccinated cases, but most Delta and Omicron studies showed no clear difference in Ct values between fully vaccinated and unvaccinated cases.
- The risk of transmission of the Omicron variant was found to be higher compared with the Delta variant, regardless of vaccination status.
- There was insufficient evidence to examine whether transmission varies by vaccine type or at different baseline community transmission levels.

###### Best quality evidence

Two studies reporting on transmission were considered high quality (Andrejko et al. 2021, Lyngse et al. 2022a), and three studies of viral load were judged to be high quality (Lyngse et al. 2022a, Migueres et al. 2022, Qassim et al. 2022).

##### Policy Implications

- The findings indicate that transmission of Omicron and Delta variants is lowest in booster-vaccinated people, followed by fully vaccinated people, with the highest rate of transmission in unvaccinated people.
- Where studies have compared transmission between different variants or sub-variants, the risk of transmission appears to be higher with Omicron than Delta, regardless of vaccination status.
- Most studies were highly heterogeneous, so caution must be used when comparing results.
- Randomised controlled trials of vaccination assessing transmission to household members or other close contacts would help to understand the true vaccine effectiveness against transmission of SARS-CoV-2. However, these would need to account for the ongoing development of new variants, which may be challenging.

##### Strength of Evidence

The new evidence consists of observational studies that are mainly good to moderate quality. However, in almost all studies there is a high risk that factors other than vaccination may have affected the findings and biased the results in either direction. The studies also varied in design, participant characteristics and SARS-CoV-2 variants reported.

## 1. New evidence on transmission of SARS-CoV-2 after COVID-19 vaccination (Table 1)

In this update, there were 6 observational studies that directly assessed the effectiveness of vaccines in reducing the risk of transmission of SARS-CoV-2 from people who had COVID-19 (index cases) to household members or close contacts (secondary cases). Four of the studies were pre-prints: Allen et al. (2022a), Jalali et al. (2022), Lyngse et al. (2022a), Sriraman et al. (2022). The six studies were appraised using the Quality Criteria Checklist (QCC) tool. Two studies were assessed as high quality (Andrejko et al. 2021, Lyngse et al. 2022a), three were of medium quality (Allen et al. 2022a, Jalali et al. 2022, Sriraman et al. 2022) and one study was of low quality (Brandal et al. 2021). Five were cohort studies (Allen et al. 2022a, Brandal et al. 2021, Jalali et al. 2022, Lyngse et al. 2022a, Sriraman et al. 2022) and one was a case-control study (Andrejko et al. 2021) One study provided data from the UK (Allen et al. 2022a), three from Europe (Brandal et al. 2021, Jalali et al. 2022, Lyngse et al. 2022a), one from Asia (Sriraman et al. 2022) and one from USA (Andrejko et al. 2021). The dates of studies being conducted ranged between January 2020 and 23 January 2022.

Two studies focussed on the Omicron variant (Brandal et al. 2021, Lyngse et al. 2022a), one study looked at the Delta variant (Sriraman et al. 2022), two studies looked at both variants (Allen et al. 2022a, Jalali et al. 2022), and one study did not report the variant, but was likely it looked at Delta as the study dates went from February 2021 to November 2021 (Andrejko et al. 2021).

The studies included people who had received COVID-19 booster vaccinations (three doses), those who were fully vaccinated (two doses), those who were partially vaccinated (one dose), and those who were unvaccinated (no vaccine doses).

Table 1 shows a summary of all transmission studies, including studies from both this update (with black text) and from the previous review (in grey text), and **<u>Supplementary Table 1</u>** shows full information for all transmission studies.

### 1.1 Evidence for the Omicron variant

#### 1.1.1 Transmission and vaccine effectiveness

There was one study from the previous review for transmission of Omicron variant SARS-CoV-2 (Lyngse et al. 2021). This study suggests that booster-vaccinated index cases transmit Omicron and Delta variant SARS-CoV-2 less than fully-vaccinated index cases, and that fully-vaccinated cases transmit less than unvaccinated index cases. There was little evidence that this was different for index cases with Omicron compared with Delta variant SARS-CoV-2. This study also suggested that there was little difference in Omicron variant SARS-CoV-2 transmission to fully vaccinated and unvaccinated household contacts, although there was less transmission to booster-vaccinated household contacts.

In addition to the study reported above, 3 studies included in this update (1 study just focused on Omicron and 2 studies looking at Omicron and Delta) looked at the difference in transmission of the Omicron variant from vaccinated and unvaccinated index cases. One study included in this update looked at the effectiveness of vaccines in reducing Omicron SARS-CoV-2 transmission. These are described in more detail below.

##### Transmission

Lyngse et al. (2022a) expanded on the study above by Lyngse et al. (2021) and assessed the transmission of two subvariants of Omicron (BA.1 and BA.2) to household members in 8,541 primary household cases in Denmark, from December 2021 to January 2022. The secondary attack rate (SAR) at seven days was estimated as 29% and 39% in households infected with Omicron BA.1 and BA.2, respectively. SAR was lowest in those index cases who were booster vaccinated compared to fully vaccinated and unvaccinated people. The study reported lower transmissibility in both BA.1 and BA.2 households when the index case was booster-vaccinated rather than fully vaccinated, with an odds ratio (OR) of 0.77 (95% confidence interval [CI]: 0.70 to 0.88) and 0.79 (95% CI: 0.64 to 0.98) for BA.1 and BA.2, respectively. There was no statistically significant difference in transmissibility in BA.1 households between unvaccinated and fully vaccinated index cases (OR: 0.93, 95% CI: 0.80 to 1.08). There was also no statistically significant difference in transmissibility between unvaccinated and fully vaccinated index cases in BA.2 households for unvaccinated index cases (OR: 1.21, 95% CI: 0.97 to 1.50) (Lyngse et al. 2022a).

Allen et al. (2022a) conducted a contact-tracing study of 51,281 SARS-CoV-2 positive people in England (13,680 with Omicron), between 5 and 11 December 2021, and found that SARs were consistently higher for Omicron than Delta for every stratum of vaccination dose for index cases or contacts.

For Delta cases, SARs were lowest among index cases who were booster-vaccinated in the household (6.2%, 95% CI: 5.3% to 7.1%) and non-household setting (3.1%, 95% CI: 2.1% to 4.2%), compared to unvaccinated people in the household setting (11.8%, 95% CI: 11.4% to 12.3%) and non-household setting (4.9%, 95% CI: 4.0% to 5.8%). The impact of index case vaccination on transmission rates for Omicron cases was considerably attenuated compared to Delta, with secondary transmission rates marginally lower for booster-vaccinated index Omicron cases (12.4%, 95% CI: 10.9% to 13.8%) compared to less than 3 doses (14.9%, 95% CI: 13.1% to 16.8%) or unvaccinated groups (15.8%, 95% CI: 14.7% to 17.0%) for all comparisons. In non-household settings, SAR was lower for Omicron cases in index cases who were vaccinated; the SAR was 8.8% (95% CI: 7.2% to 10.4%) for those who were unvaccinated, 7.5% (95% CI: 6.9% to 8.0%) for those who were fully vaccinated and 7.1% (95% CI: 5.7% to 8.4%) for those who were booster vaccinated (Allen et al. 2022a).

In addition, the proportion of index cases resulting in residential clustering was twice as high for Omicron (16.1%) compared to Delta (7.3%) (Allen et al. 2022a). The risk ratio of household clustering found that the overall risk ratio was 3.54 (95% CI: 3.29 to 3.81) for Omicron compared to Delta variants. Furthermore, for each of vaccination status there was an increased risk of household clustering for Omicron compared to Delta variants, most notably among index cases who were ≥14 days post their third vaccine dose, with a risk ratio of household clustering of 6.81 comparing Omicron and Delta variants (95% CI: 4.91 to 9.46) (Allen et al. 2022a).

Another contact-tracing study by Jalali et al. (2022), of 1,122 index cases and 2,169 household contacts in Norway, found an overall higher household ten-day SAR for the Omicron variant (51%, 95% CI: 48% to 54%) compared to the Delta variant (36%, 95% CI: 33% to 40%), with a relative risk of 1.41 (95% CI: 1.27 to 1.56). Index cases who were booster vaccinated were found to have a considerably higher risk (relative risk: 4.34; 95% CI: 1.52 to 25.16) of transmitting SARS-CoV-2 to their household contacts with Omicron compared to Delta. A similar trend was observed when the index case was unvaccinated or fully vaccinated, although the relative risks were lower (unvaccinated relative risk: 1.51, 95% CI: 1.30 to 1.77; fully vaccinated relative risk: 1.44, 95% CI: 1.24 to 1.70). There was not a significant increase in transmissibility in Omicron index cases who were partially vaccinated, compared to Delta (relative risk: 1.05, 95% CI: 0.76 to 1.52). Fully vaccinated index cases had a similar risk as unvaccinated index cases in Omicron transmission to their adult household members (relative risk: 1.04; 95% CI: 0.79 to 1.49). The same pattern was observed for the booster-vaccinated versus unvaccinated index cases (relative risk: 0.99; 95% CI: 0.68 to 1.49). In contrast, booster-vaccinated index cases with Delta had an 80% lower risk of transmission (0.18; 95% CI: 0.01 to 0.70) relative to unvaccinated index cases (Jalali et al. 2022).

##### Vaccine effectiveness

The effectiveness of vaccines in reducing infection for booster-vaccinated adult contacts was lower for Omicron (45%; 95% CI: 26% to 57%) compared to Delta (65%; 95% CI: 42% to 80%) but higher than for fully vaccinated adults (Jalali et al. 2022).

Brandal et al. (2021) conducted a cohort study of 110 people in Norway who attended a party, including someone who had recently travelled to South Africa and tested positive for Omicron (following a targeted exploration due to travel) in the days following the party. Most of the attendees (96%) were fully vaccinated with two vaccine doses (but no booster doses). The cohort was tested for Omicron in the 17 days following the party: 59% were confirmed cases and 14% were probable cases of Omicron. The total attack rate for the Omicron variant was 74% (Brandal et al. 2021). Although this study didn’t compare vaccinated people with unvaccinated, or levels of vaccination, it was included as it provided transmission data for the Omicron variant in fully vaccinated people.

#### 1.1.2 Summary of the evidence for Omicron transmission and vaccine effectiveness

Evidence from the current review supports the findings of the study included in the previous review by Lyngse et al. (2021) that booster-vaccinated index cases transmit Omicron and Delta variant SARS-CoV-2 less than fully vaccinated index cases, who transmit less than unvaccinated index cases (Allen et al. 2022a, Brandal et al. 2021, Jalali et al. 2022, Lyngse et al. 2022a). Furthermore, one of the studies included in this review found that the same is true for both subvariants of Omicron (BA. and BA.2) (Lyngse et al. 2022a).

The study by Lyngse et al. (2021) in the previous review and the studies identified in this review found evidence that the risk of transmission for index cases with Omicron was higher compared with Delta variant SARS-CoV-2, in those of all vaccination exposure statuses (Allen et al. 2022a, Jalali et al. 2022, Lyngse et al. 2021).

The evidence included in this review and the previous review also suggests that transmission was significantly less likely to contacts who had received booster vaccines compared to contacts who were fully vaccinated or partially vaccinated (Allen et al. 2022a, Lyngse et al. 2021).

### 1.2 Evidence for the Delta variant

There were 10 studies in the previous review (Allen et al. 2022b, Clifford et al. 2021, de Gier et al. 2021a, Eyre et al. 2022, Hsu et al. 2021, Kang et al. 2022, Lyngse et al. 2021, Martinez-Baz et al. 2021, Ng et al. 2021, Singanayagam et al. 2021) which suggested fully vaccinated cases transmitted Delta variant SARS-CoV-2 less than unvaccinated cases. Two of these studies suggested that vaccine effectiveness against transmission of the Delta variant dropped substantially over time, though no studies in the update assessed this. Additionally, evidence from 9 studies in the previous review (Clifford et al. 2021, Eyre et al. 2022, Hsu et al. 2021, Lyngse et al. 2021, Lyngse et al. 2022b, Martinez-Baz et al. 2021, Ng et al. 2021, Singanayagam et al. 2021, Yi et al. 2022) suggested that index cases typically transmitted SARS-CoV-2 to fully vaccinated contacts less than unvaccinated contacts.

Two studies included in this update looked at the difference in transmission of the Delta variant SARS-CoV-2 alone from vaccinated and unvaccinated index cases (Andrejko et al. 2021, Sriraman et al. 2022).

Andrejko et al. (2021) found that the odds of cases status were lower for both partially (OR: 0.30, 95% CI: 0.15 to 0.43) and fully vaccinated (OR: 0.25, 95% CI: 0.15 to 0.43) participants relative to unvaccinated participants. The study also reported that of the odds ratio of participants with case status was 3.02 (95% Cl: 1.75 to 5.22) when high-risk exposures occurred with household members (versus other contacts), 2.10 (95% CI: (1.05 to 4.21) when exposures occurred indoors (versus outdoors), and 2.15 (95% CI: 1.27 to 3.67) when exposures lasted ≥3 hours (versus shorter durations) among unvaccinated and partially-vaccinated individuals; excess risk associated with such exposures was mitigated among fully-vaccinated individuals.

A prospective, observational study of 92 index cases in India (pre-print) by Sriraman et al. (2022) reported that the median household infection rate (HHR: defined as the number of confirmed positive members in the household at enrolment + number of new positive members at follow-up + number of new positive members at final telephone follow-up)/total number of individuals living in the same household) was significantly higher in people partially vaccinated with AstraZeneca (median HHR: 67%) and fully vaccinated with AstraZeneca (median HHR: 88%) compared to the unvaccinated (median HHR: 20%) (Sriraman et al. 2022).

#### 1.2.1 Summary of the evidence for Delta transmission

Overall, evidence from 10 studies from the previous review (Allen et al. 2022b, Clifford et al. 2021, de Gier et al. 2021a, Eyre et al. 2022, Hsu et al. 2021, Kang et al. 2022, Lyngse et al. 2021, Martinez-Baz et al. 2021, Ng et al. 2021, Singanayagam et al. 2021) and one of the studies in this review (Andrejko et al. 2021) suggested fully vaccinated cases transmitted Delta variant SARS-CoV-2 less than unvaccinated cases. However, the one new study identified in this review did not find a significant difference in the risk of transmission amongst vaccinated and unvaccinated people with the Delta variant (Sriraman et al. 2022).

Two studies from the previous review suggested that vaccine effectiveness against transmission of the Delta variant dropped substantially over time. Additionally, evidence from 9 studies from the previous review (Clifford et al. 2021, Eyre et al. 2022, Hsu et al. 2021, Lyngse et al. 2021, Lyngse et al. 2022b, Martinez-Baz et al. 2021, Ng et al. 2021, Singanayagam et al. 2021, Yi et al. 2022) suggested that index cases typically transmitted SARS-CoV-2 to fully vaccinated contacts less than unvaccinated contacts.

### 1.3 Evidence for pre-Delta variants

Evidence from 15 studies from the previous review (Allen et al. 2021, Braeye et al. 2021, de Gier et al. 2021b, Harris et al. 2021a, Harris et al. 2021b, Layan et al. 2022, Meyer et al. 2021, Prunas et al. 2022, Shah et al. 2021, Allen et al. 2022b, Bobdey et al. 2021, Clifford et al. 2021, Hsu et al. 2021, Martinez-Baz et al. 2021, Ng et al. 2021) suggested that fully vaccinated index cases transmitted pre-Delta variant and Wild-type SARS-CoV-2 to their contacts less than unvaccinated index cases, and this reduction was substantial (e.g. >50% reduction in transmission) in many studies. No new evidence was identified in this review.

## 2. New evidence on viral load in those who develop COVID-19 infection after being vaccinated (Table 2)

In this update, we identified 11 additional observational studies that compared viral loads (predominantly using Ct values) between vaccinated and unvaccinated COVID-19 cases. Six were pre-prints (Boucau et al. 2022, Fall et al. 2022, Kislaya et al. 2022a, Lyngse et al. 2022a, Qassim et al. 2022, Sriraman et al. 2022). Of 11 studies, three studies were judged to be high quality (Lyngse et al. 2022a, Migueres et al. 2022, Qassim et al. 2022), six of medium quality (Accorsi et al. 2022, Boucau et al. 2022, Fall et al. 2022, Kislaya et al. 2022a, Sriraman et al. 2022, Williams et al. 2021) and two were of low quality (Kissler et al. 2021, Barbosa et al. 2022). Of these, nine were cohort studies (Barbosa et al. 2022, Boucau et al. 2022, Fall et al. 2022, Kissler et al. 2021, Lyngse et al. 2022a, Migueres et al. 2022, Qassim et al. 2022, Sriraman et al. 2022, Williams et al. 2021), and 2 were case-control studies (Accorsi et al. 2022, Kislaya et al. 2022a). One of the studies provided data from the UK (Williams et al. 2021), 4 from the USA (Accorsi et al. 2022, Boucau et al. 2022, Fall et al. 2022, Kissler et al. 2021), 3 from Europe (Kislaya et al. 2022a, Lyngse et al. 2022a, Migueres et al. 2022), 2 from Asia (Qassim et al. 2022, Sriraman et al. 2022) and one from Brazil (Barbosa et al. 2022).

All studies were conducted between November 2020 and February 2022.

Three studies looked at the Omicron variant (Fall et al. 2022, Lyngse et al. 2022a, Qassim et al. 2022), three looked at both Omicron and Delta variants (Accorsi et al. 2022, Boucau et al. 2022, Kislaya et al. 2022a), one looked at the Delta variant (Sriraman et al. 2022), two looked at both Delta and Alpha variants (Kissler et al. 2021, Migueres et al. 2022), one looked at the Alpha variant (Williams et al. 2021), and one study included data without reporting the variant (Sriraman et al. 2022).

**Table 2** shows a summary of all viral load studies, including studies from both this update (with black text) and from the previous review (in grey text), and **Supplementary Table 2** shows full information for all viral load studies.

### 2.1 Evidence for the Omicron variant

Two studies included in the previous review compared the viral loads of Omicron and Delta variant cases (Puhach et al. 2022, Lyngse et al. 2021), though neither study reported the difference in Ct value between vaccinated and unvaccinated cases.

Six studies included in this update compared the viral load of Omicron in vaccinated and unvaccinated people (Accorsi et al. 2022, Boucau et al. 2022, Fall et al. 2022, Kislaya et al. 2022a, Lyngse et al. 2022a, Qassim et al. 2022).

#### 2.1.1 Viral load (Ct values)

A retrospective test-negative case-control analysis conducted in the USA measured median Ct values for three viral genes, stratified by variant and vaccination status, in 23,391 people with a positive SARS-CoV-2 test (Accorsi et al. 2022). Median Ct values were significantly higher (q-value < 0.001), indicating lower viral load, in cases who were booster vaccinated versus those who were fully vaccinated, for both Omicron and Delta (Omicron N gene: 19.35 versus 18.52; Omicron ORF1ab gene: 19.25 versus 18.40; Delta N gene: 19.07 versus 17.52; Delta ORF1ab gene: 18.70 versus 17.28; Delta S gene: 23.62 versus 20.24). Among Omicron cases, median Ct values were slightly higher, indicating lower viral load, in samples from booster-vaccinated people versus unvaccinated people for the ORF1ab gene (19.25 versus 18.58) but not for the N gene (19.35 versus 18.71). Among Delta cases, median Ct values of the N, ORF1ab, and S genes were slightly higher (indicating lower viral load) in samples from people who were booster vaccinated versus unvaccinated (N gene: 19.07 versus 18.28; ORF1ab gene: 18.70 versus 17.84; S gene: 23.62 versus 19.58) (Accorsi et al. 2022).

A pre-print by Fall et al. (2022) reported no statistically significant difference when Ct values were compared between vaccinated and unvaccinated patients from: Omicron vaccinated groups (n = 235), Omicron unvaccinated (n = 170), Delta vaccinated (n = 240), and Delta unvaccinated (n = 411) groups (Fall et al. 2022).

Kislaya et al. (2022a) conducted a case-case study in Portugal of 4,898 people aged 12 years and older with Omicron and 8,245 people aged 12 years and older with Delta variants. For patients aged 50 years and older, of which 18% were booster vaccinated, no statistically significant differences in mean Ct values by variant or vaccination status were observed. They did not find a significant mean difference in Ct values between these two variants for unvaccinated (-0.002, 95% CI: -0.6 to 0.6), partially vaccinated (-1.1, 95% CI: -2.6 to 0.36) or fully vaccinated people (-0.01, 95% CI: -0.2 to 0.2).

##### Viral load (Ct values): Omicron subvariants

A retrospective cohort study by Lyngse et al. (2022a) of 8,541 primary household cases in Denmark assessed the transmission of two subvariants of Omicron (BA.1 and BA.2) to household members. The study reported that the distribution of sample Ct values for unvaccinated index cases was higher overall for BA.2 cases than for BA.1 cases. This was not the case for fully vaccinated and booster-vaccinated individuals, where the distribution appeared to be similar.

A similar cross-sectional study of 156,202 people in Qatar also investigated the effects of vaccination on the Ct values of Omicron subvariants BA.1 and BA.2 (Qassim et al. 2022). They found that compared to BA.1, BA.2 was associated with a lower Ct value (-3.53, 95% CI: -3.46 to -3.60). Ct values decreased with time since second and third vaccinations, following the established pattern of waning vaccine effectiveness. Ct values were highest for those who received their boosters in the month preceding the RT-qPCR test: 0.86 cycles (95% CI: 0.72 to 1.00) higher than for unvaccinated persons (Qassim et al. 2022).

#### 2.1.2 Viral load (Ct values, time since vaccination)

Considering time since completion of primary vaccination, Kislaya et al. (2022a) observed statistically significant differences between Delta and Omicron cases only for cases with less than 113 days since complete primary vaccination (mean difference: -0.70; 95% CI: -1.13 to -0.28), indicating slightly lower Ct (higher viral loads) in Omicron cases. There were no statistically significant differences between Delta and Omicron for cases more than 113 days since complete primary vaccination (Kislaya et al. 2022a).

In the pre-print mentioned above by Fall et al. (2022), when the Ct analysis was correlated to the days from the onset of symptoms for symptomatic patients, no significant differences were detected between Omicron and Delta, considering fully vaccinated, boosted, and unvaccinated cases separately. The authors concluded that there were no significant differences in viral RNA loads between Omicron and Delta infected individuals (Fall et al. 2022).

#### 2.1.3 Viral load (time to viral clearance)

A pre-print by Boucau et al. (2022) reported on how Omicron and Delta variants and vaccination status impact shedding of viable virus in an American longitudinal cohort study of 56 non-severely symptomatic patients with COVID-19. Viral load decay and time to negative PCR (time to PCR conversion) did not differ between participants infected with Omicron versus Delta (hazard ratio [HR]: 0.85, 95% CI: 0.44 to 1.61). Duration of shedding of viable virus, as measured by time to culture conversion, was also similar by variant (HR: 0.86, 95% CI: 0.47 to 1.58), with a median time to culture conversion of 6 days in both groups (IQR 4 to 8 days). In the overall cohort, there was no significant difference in time to PCR conversion (p = 0.08) or culture conversion (p = 0.57) by vaccination status.

#### 2.1.4 Summary of the evidence for Omicron viral load

Evidence from 2 studies in the previous review (Lyngse et al. 2021, Puhach et al. 2022) suggests there is little difference between the Ct values and genome copy numbers of Omicron and Delta variant SARS-CoV-2 cases, though infectious viral loads may be smaller in Omicron cases. This is supported by 3 studies in this current review (Fall et al. 2022, Kislaya et al. 2022a, Boucau et al. 2022). However, a large study found that median Ct values were significantly higher in cases with booster doses versus cases who were fully vaccinated, for both Omicron and Delta, and between individuals who were booster vaccinated and unvaccinated (Accorsi et al. 2022), and another study found slightly higher viral loads in Omicron compared to Delta for those cases with less than 113 days since complete primary vaccination (Kislaya et al. 2022a).

Two studies in this review reported Ct values in the Omicron subtypes BA.1 and BA.2 (Lyngse et al. 2022a, Qassim et al. 2022). One study found that viral load was overall higher for BA.2 cases than for BA.1 cases in unvaccinated people, but this was not the case for fully vaccinated and booster-vaccinated individuals (Lyngse et al. 2022a). The other study also reported viral load as being higher for BA.2 cases and that Ct values were highest for those who received their boosters in the month preceding the PCR test compared to unvaccinated cases (Qassim et al. 2022).

### 2.2 Evidence for the Delta variant

#### 2.2.1 Viral load (Ct values)

Evidence from 25 studies from the previous review (Acharya et al. 2021, Chia et al. 2021, Christensen et al. 2022, Elliott et al. 2021, Eyre et al. 2022, Griffin et al. 2021, Hirotsu et al. 2021, Hsu et al. 2021, Kale et al. 2021, Kang et al. 2022, Kerwin et al. 2021, Kislaya et al. 2022b, Levine-Tiefenbrun et al. 2021a, Levine-Tiefenbrun et al. 2022, Luo et al. 2021a, Lyngse et al. 2022b, Magalis et al. 2021, Pouwels et al. 2021, Puhach et al. 2022, Riemersma et al. 2021, Salvatore et al. 2021, Servellita et al. 2022, Siddle et al. 2022, Singanayagam et al. 2021, Yi et al. 2022) suggests mixed evidence for a difference in viral load between fully vaccinated and unvaccinated cases, with 17 studies suggesting no difference and 8 studies suggesting higher Ct values (lower viral load) in fully vaccinated cases.

A further three studies have been included in this update, which report on Ct levels in vaccinated and unvaccinated Delta cases (Barbosa et al. 2022, Migueres et al. 2022, Sriraman et al. 2022).

A study of 1,059 vaccinated healthcare workers in Brazil was published in a letter by Barbosa et al. (2022) and did not observe a significant difference in the viral load regardless of vaccine type or number of doses (Barbosa et al. 2022).

A prospective study in France by Migueres et al. (2022) reported that the nasopharyngeal viral loads of unvaccinated patients infected with the Delta variant were greater than in fully vaccinated patients (median 7.1 versus 6.64, p < 0.001). The difference was similar for the Alpha variant but not significant (the authors suggest that this is due to the small sample size).

Another prospective study of 92 people with COVID-19 in India (pre-print) by (Sriraman et al. 2022) found no significant difference in the expelling pattern (high-risk emitters [people expelling >1000 viral copy numbers in 30 minutes], medium-risk emitters [A viral copy number of 100-999] and low-risk emitters [< 100 copy numbers] of SARS-CoV-2) between partial, full and unvaccinated individuals, suggesting similar transmission risk. In addition, the viral load data showed that only patients in the partially and fully vaccinated AstraZeneca groups had a statistically significant reduction in viral load at follow-up (8 to 12 days after COVID-19 diagnosis), compared to other vaccines and unvaccinated cases.

Overall, evidence from 28 studies (25 studies from the previous review (Acharya et al. 2021, Chia et al. 2021, Christensen et al. 2022, Elliott et al. 2021, Eyre et al. 2022, Griffin et al. 2021, Hirotsu et al. 2021, Hsu et al. 2021, Kale et al. 2021, Kang et al. 2022, Kerwin et al. 2021, Kislaya et al. 2022b, Levine-Tiefenbrun et al. 2021a, Levine-Tiefenbrun et al. 2022, Luo et al. 2021a, Lyngse et al. 2022b, Magalis et al. 2021, Pouwels et al. 2021, Puhach et al. 2022, Riemersma et al. 2021, Salvatore et al. 2021, Servellita et al. 2022, Siddle et al. 2022, Singanayagam et al. 2021, Yi et al. 2022), and three studies from this update (Barbosa et al. 2022, Migueres et al. 2022, Sriraman et al. 2022), suggests mixed evidence for a difference in viral load between fully vaccinated and unvaccinated cases, with 17 studies suggesting no difference and 11 studies suggesting higher Ct values (lower viral load) in fully vaccinated cases.

#### 2.2.2 Viral load (Ct values, time since vaccination)

In the previous preview, evidence from 2 studies (Levine-Tiefenbrun et al. 2021a), (Levine-Tiefenbrun et al. 2022) suggested that although Ct values are higher in fully and booster-vaccinated cases compared with unvaccinated cases (lower viral load) soon after vaccination, this difference drops quickly, with Ct values becoming similar between 61 and 120 days after vaccination.

#### 2.2.3 Viral load (time to viral clearance)

Evidence from 4 studies in the previous review (Hagan et al. 2021, Kang et al. 2022, Salvatore et al. 2021, Singanayagam et al. 2021) suggested that there was not a large or statistically significant difference in the time to viral clearance between fully vaccinated and unvaccinated cases.

Two studies investigating viral clearance were included in this update (Kissler et al. 2021, Sriraman et al. 2022).

SARS-CoV-2 viral dynamics were investigated in a prospective, longitudinal study, published in a letter by (Kissler et al. 2021), with 37 vaccinated and 136 unvaccinated people. Vaccinated individuals exhibited faster clearance compared to unvaccinated (mean: 5.5 days [95% CI: 4.6 to 6.5] versus 7.5 days [95% CI: 6.8 to 8.2]).

A prospective, observational study of 92 people with COVID-19 in India (pre-print) by Sriraman et al. (2022) found that among the vaccinated groups, the patients in the fully vaccinated Covaxin group (Bharat Biotech) had significantly higher mask positivity (proportion of patients expelling the virus) at 8 days (90%) compared to the partially and fully AstraZeneca vaccinated groups, suggesting that Covaxin patients may be clearing the virus slower than the AstraZeneca vaccinated groups and unvaccinated.

Overall, evidence from four studies in the previous review (Hagan et al. 2021, Kang et al. 2022, Salvatore et al. 2021, Singanayagam et al. 2021) did not suggest large or statistically significant differences in the time to viral clearance between fully vaccinated and unvaccinated cases. However, evidence from the 2 studies in this update suggest that vaccinated individuals exhibit faster viral clearance compared to unvaccinated (Kissler et al. 2021, Sriraman et al. 2022).

#### 2.2.4 Infectious viral load (cytopathic effects)

Evidence from 6 studies from the previous review (Hagan et al. 2021, Luo et al. 2021a, Peña-Hernández et al. 2022, Puhach et al. 2022, Riemersma et al. 2021, Salvatore et al. 2021), suggested there was little difference in cytopathic effects between fully vaccinated and unvaccinated cases. No new evidence was identified in this review for this outcome.

### 2.3 Evidence for pre-Delta variants

From 27 studies from the previous review (Adamson et al. 2021, Abu-Raddad et al. 2022, Bailly et al. 2022, Blanquart et al. 2021, Boschi et al. 2021, Brunner-Ziegler et al. 2022, Costa et al. 2022, Emary et al. 2021, Eyre et al. 2022, Griffin et al. 2021, Hsu et al. 2021, Ioannou et al. 2021, Jacobson et al. 2022, Kislaya et al. 2022b, Kolobukhina et al. 2021, Lumley et al. 2021, Luo et al. 2021a, McEllistrem et al. 2021, Mostafa et al. 2021, Muhsen et al. 2021, Pajon et al. 2021, Pouwels et al. 2021, Regev-Yochay et al. 2021, Servellita et al. 2022, Smith et al. 2022, Tande et al. 2021, Thompson et al. 2021), there was evidence suggesting fully vaccinated cases had higher Ct values than unvaccinated cases (suggesting a lower viral load).

One study was identified in this update, which involved a retrospective analysis of test trending data of arrivals into the UK, and reported mean Ct values of positive cases from vaccinated (n = 274) and unvaccinated (n = 2,417) individuals as being 25.42 and 25.55, respectively, indicating no significant difference in the viral loads between these two groups (Williams et al. 2021).

## 3. Inequalities

There was little evidence availably to explore inequalities through variations across populations and subgroups, for example cultural variations or differences between ethnic, social, or vulnerable groups, either in the previous review or this update. As such, it was not possible to examine inequalities in this report.

## 4. Limitations

The source of evidence in this review included peer-reviewed and pre-print articles. We did not conduct an extensive search of other sources (such as websites of public health organisations).

All studies were observational, comparing people who were vaccinated with those who were not. Therefore, there is a high risk in all studies that factors other than vaccination affected the results. This includes factors such as behaviour (including test seeking behaviour and behaviours likely to alter the risk of SARS-CoV-2 transmission), individual characteristics (such as age, sex, and deprivation), and COVID-19 characteristics (such as variant and symptom status). Partly due to this heterogeneity and partly due to a lack of evidence, we were unable to assess how the risk of onward transmission varied with different vaccine types and baseline community transmission levels. Few studies (4 of 43 in the previous review, 5 of 17 in this update) were rated as high quality using the QCC tool, largely because few studies accounted for these risks well.

Most studies were heterogeneous, in terms of their location, prevalence of COVID-19 in the community, prevalence of past infections, dominant variant, background mitigations in place to limit transmission (including both local restrictions and personal protective measures), vaccination status of contacts, and availability of the vaccine to different groups, as well as the demographics of the index cases, household members and other close contacts. This makes direct comparison between studies and specific vaccines difficult. However, there were four studies offering medium to high quality evidence from the UK for the Delta variant (Allen et al. 2022a, Allen et al. 2022b, Eyre et al. 2022, Williams et al. 2021).

As with all reviews, the evidence identified may be subject to publication bias, whereby null or negative results are less likely to have been published by the authors. Ten of the 25 studies identified in this update were pre-prints and should be treated with caution as they have not been peer reviewed or subject to publishing standards and may be subject to change. This is in addition to 19 pre-prints or non-peer reviewed reports of the 43 studies identified in the previous review, although 2 of these have since been published (Gazit et al. 2021a, Luo et al. 2021b). In addition, our rapid review is limited by the fact that we are reviewing evidence from an emerging field that spans less than 1 year, and only 2 months for the currently dominant Omicron variant. Studies conducted in the COVID-19 context are conducted at pace with the aim to provide evidence in a timely manner, which sometimes impacts on the quality of the studies, both in term of design (especially limited statistical analyses) and reporting (insufficient detail). There is currently little evidence for the recently identified Omicron variant (Lyngse et al. 2021, Puhach et al. 2022).

## 5. Conclusion

The main conclusions from the previous review were not changed from the inclusion of 17 additional studies in this update. There was evidence that fully and booster-vaccinated cases transmit SARS-CoV-2 less than unvaccinated cases, particularly for pre-Delta and Wild-type variants, and there was evidence suggesting that this difference is reduced with increasing time after the vaccine dose, particularly for the Delta and Omicron variants. The results from viral load studies are broadly supportive of these results, with most pre-Delta studies showing fully vaccinated cases have larger Ct values or lower viral loads than unvaccinated cases, and most Delta and Omicron studies showing no clear difference in Ct values between fully vaccinated and unvaccinated cases.

The transmission studies that reported Omicron data included in this update suggested that fully- and booster-vaccinated index cases can transmit both Omicron and Delta variant SARS-CoV-2 less than unvaccinated index cases, with the risk of transmission of Omicron being higher compared with the Delta variant SARS-CoV-2.

In almost all included studies (transmission and viral load) there is a high risk that factors other than vaccination may have affected the results, which may have biased the results in either direction. Most studies were also highly heterogeneous, so caution must be used when comparing results between different studies. Partly because of this heterogeneity, there was insufficient evidence to examine whether transmission varies by vaccine type or at different baseline community transmission levels.

## 6. Research needed

Randomised controlled trials of vaccination assessing transmission to household members or other close contacts would help us to understand the true vaccine effectiveness against transmission of SARS-CoV-2, and from the previous review, we are aware of 2 ongoing RCTs, 1 in the US (NCT04811664, estimated publication date December 2021) and 1 in the UK (NCT04750356, estimated publication date December 2024), that could help estimate this. A cross-sectional school-based study to investigate SARS-CoV-2 transmission dynamics and the impact of different variants among confirmed cases and classmates is also ongoing (Bordas et al. 2022). These studies are described in **Supplementary** Table 3. The results of these trials have not yet been published as of 25 May 2022.

## Data Availability

All data produced in the present study are available upon reasonable request to the authors

## 7. Acknowledgments

We would like to thank colleagues within the Public Health Advice, Guidance and Expertise function who either reviewed or input into aspects of the previous or this updated review, especially Helen McAuslane and Mario Aramouni. Also, thanks to Welsh Government and Public Health Wales stakeholders Simon Rolfe and Catherine Moore for their support of this work. We would also like to thank Sean Harrison and Rachel Clarke from the UK Health Security Agency (UKHSA), who had considerable contributions to the report.

## 8. Summary Tables

**Table 1.**
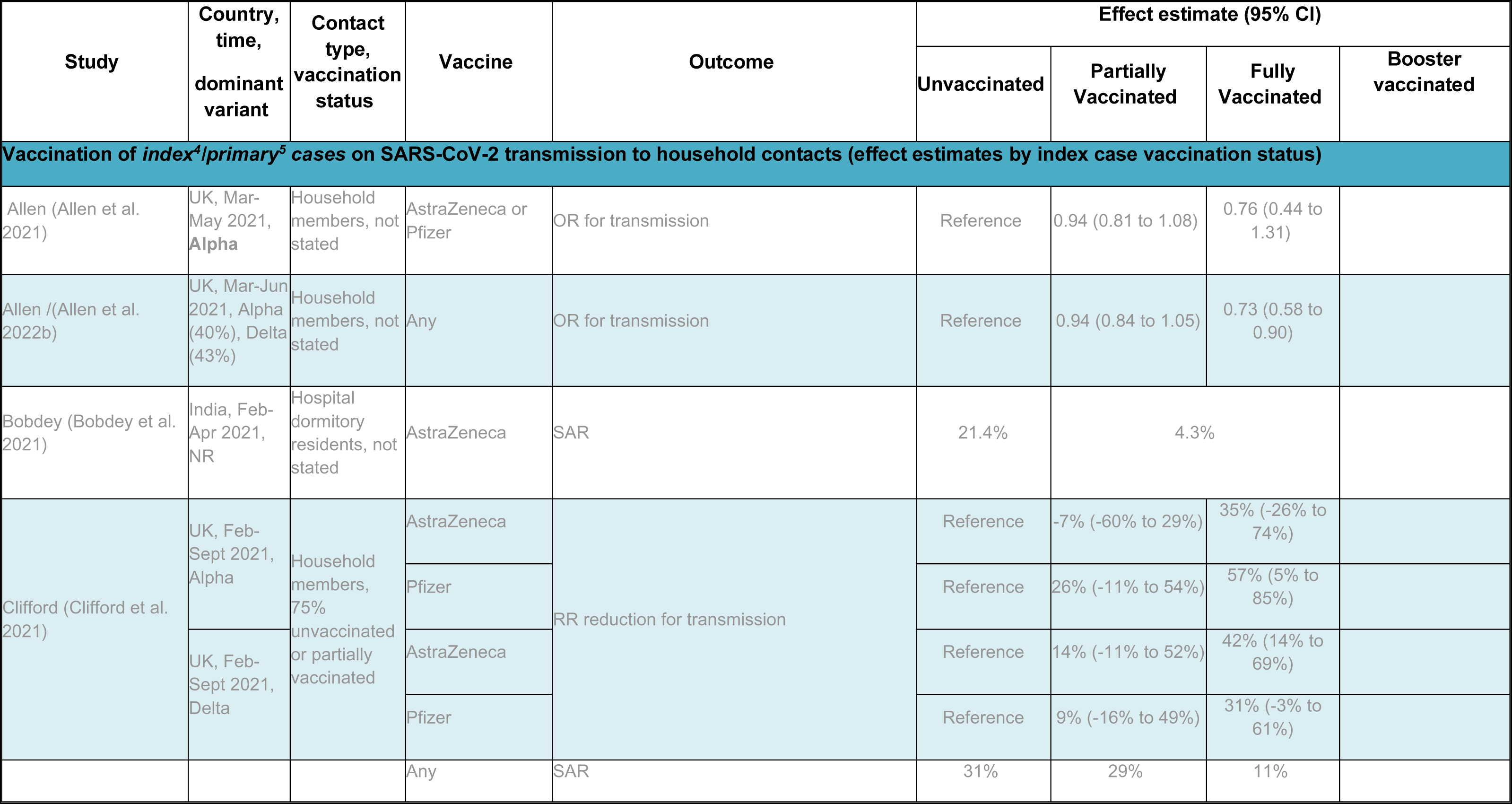
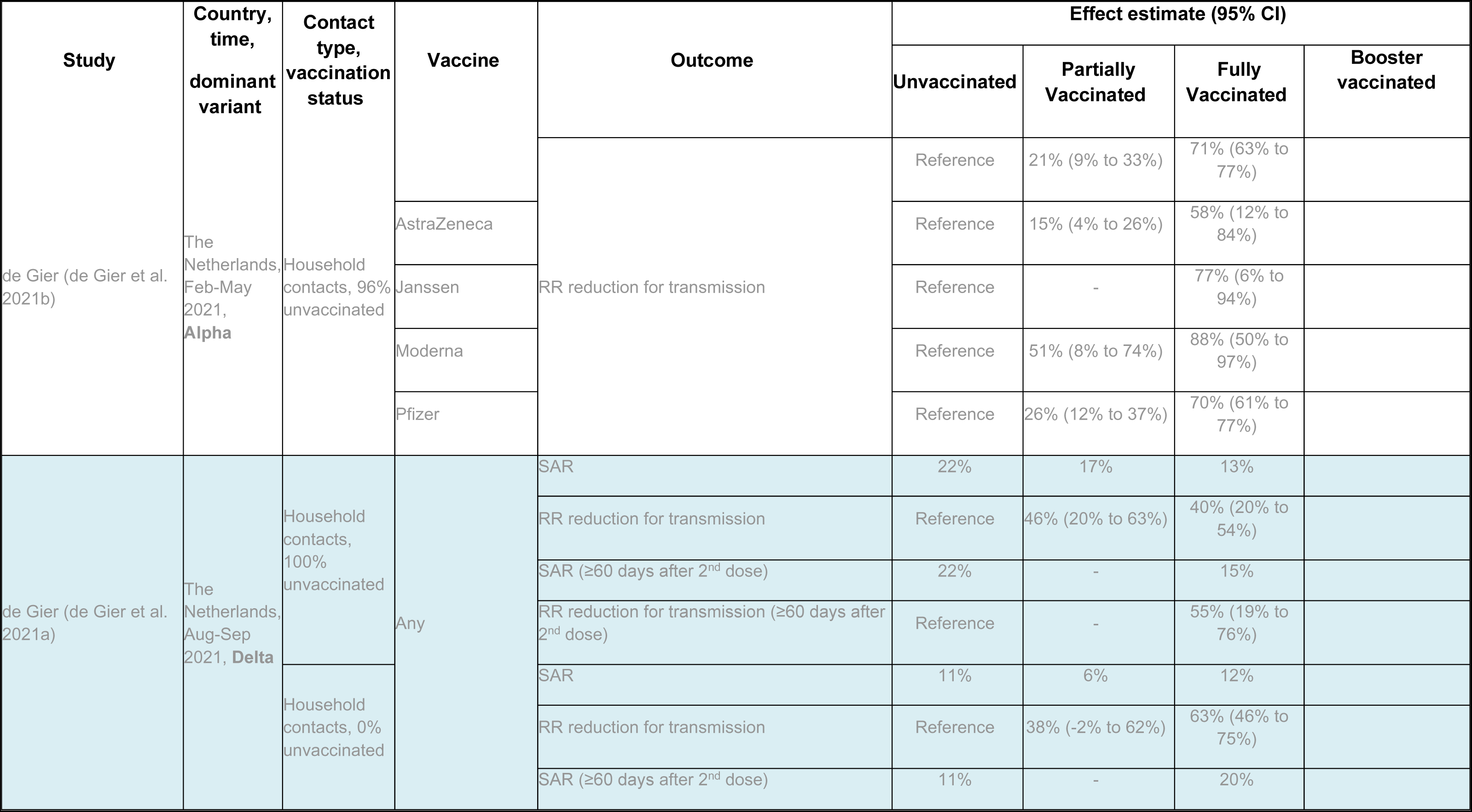
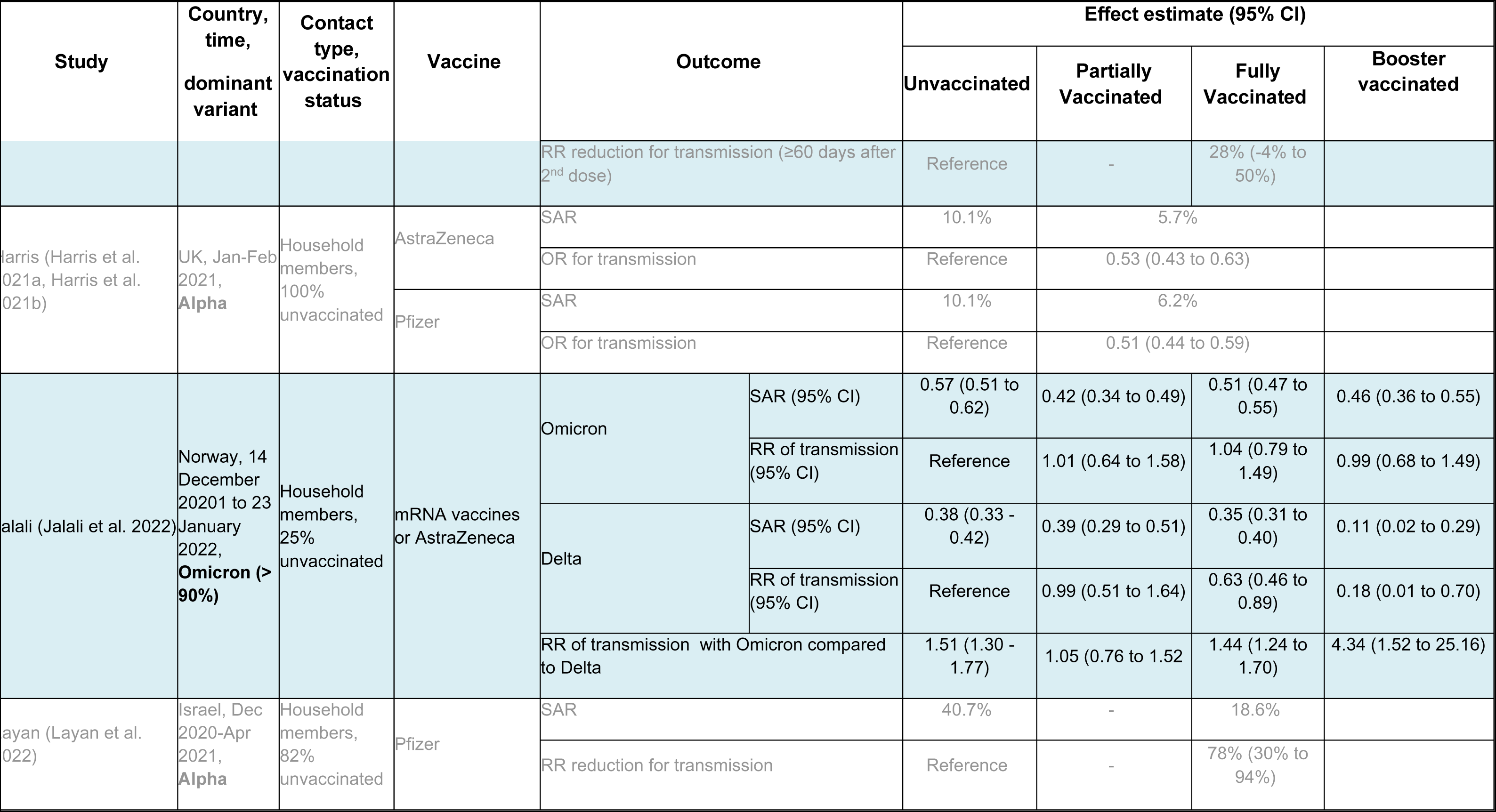
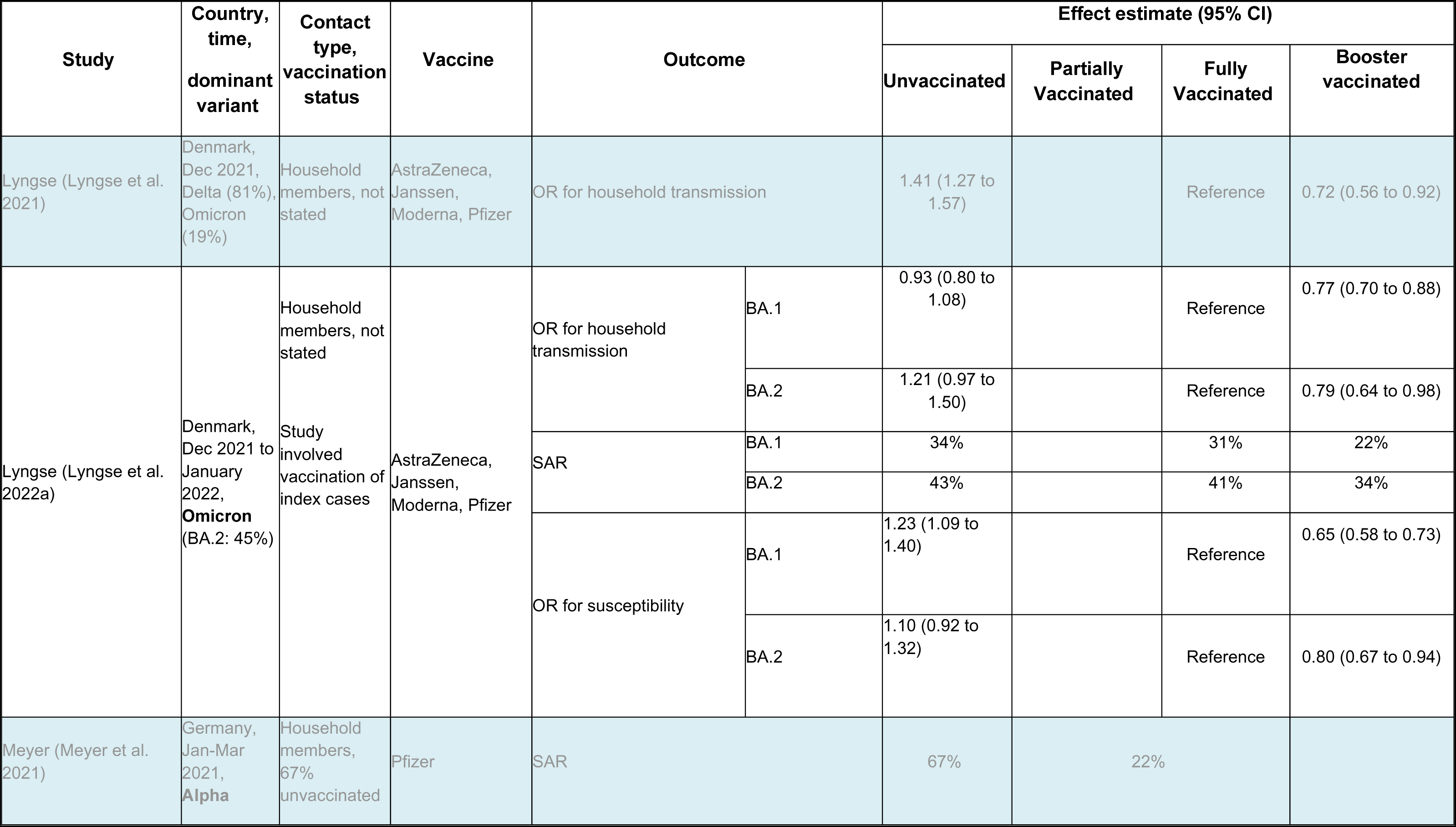
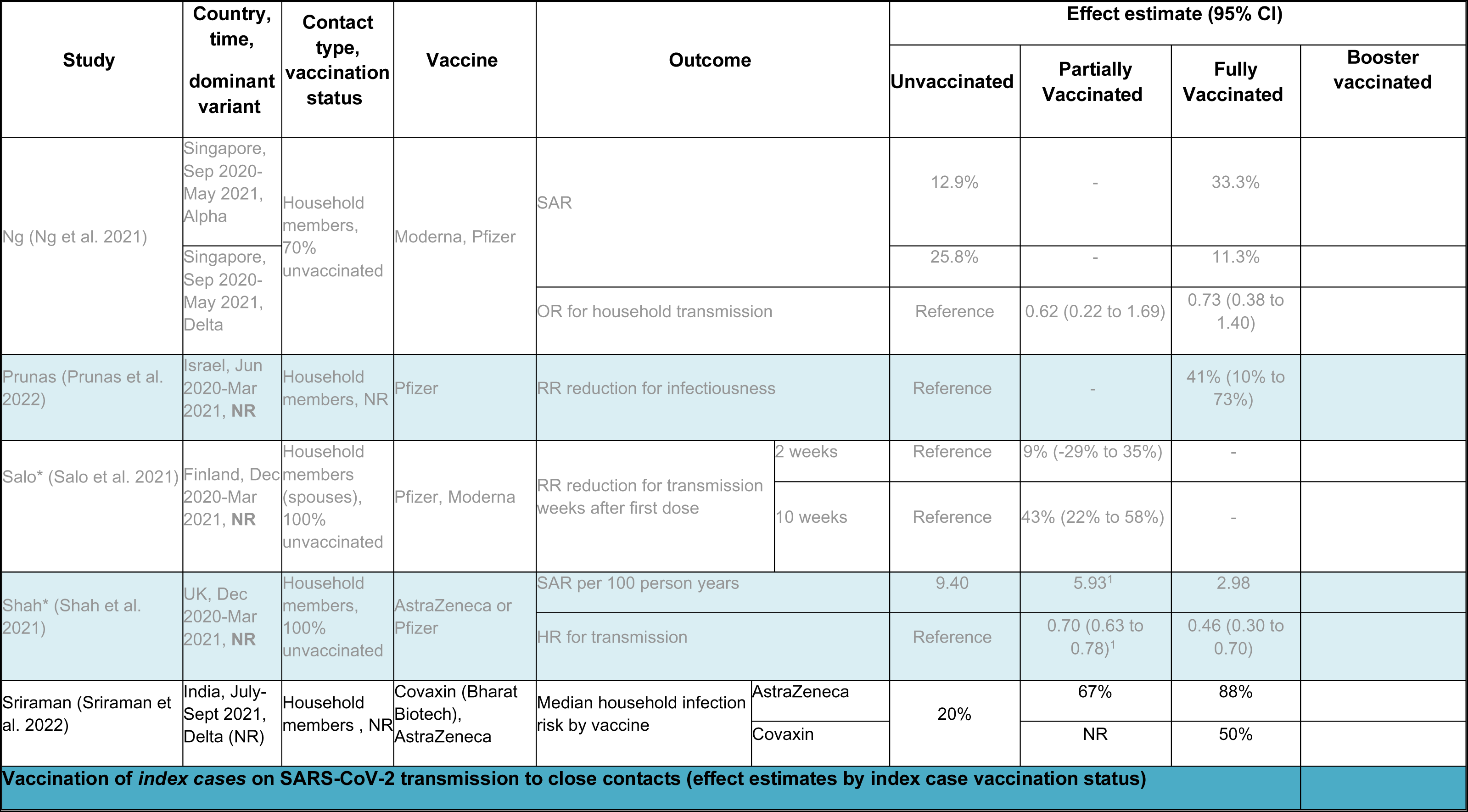
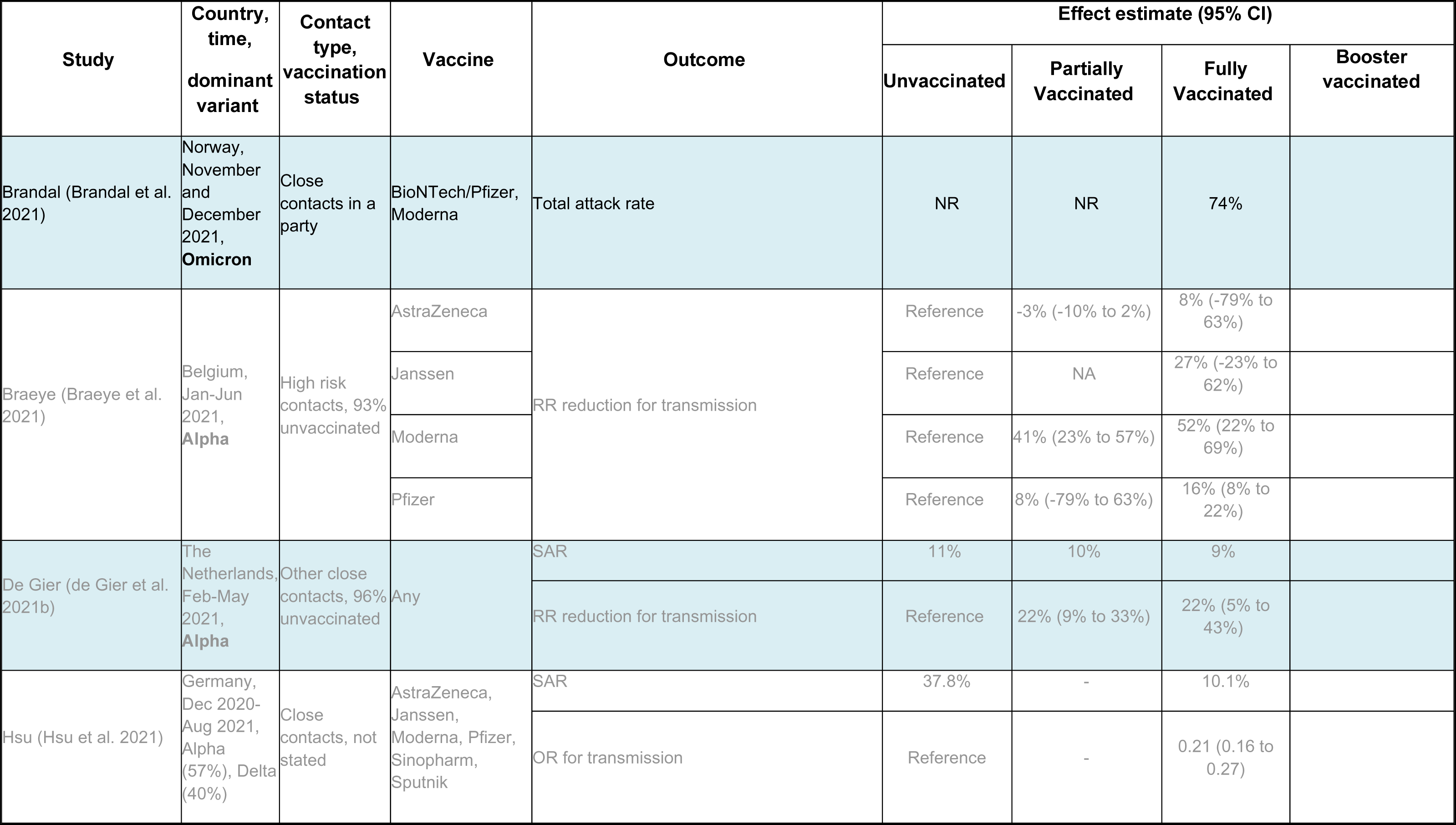
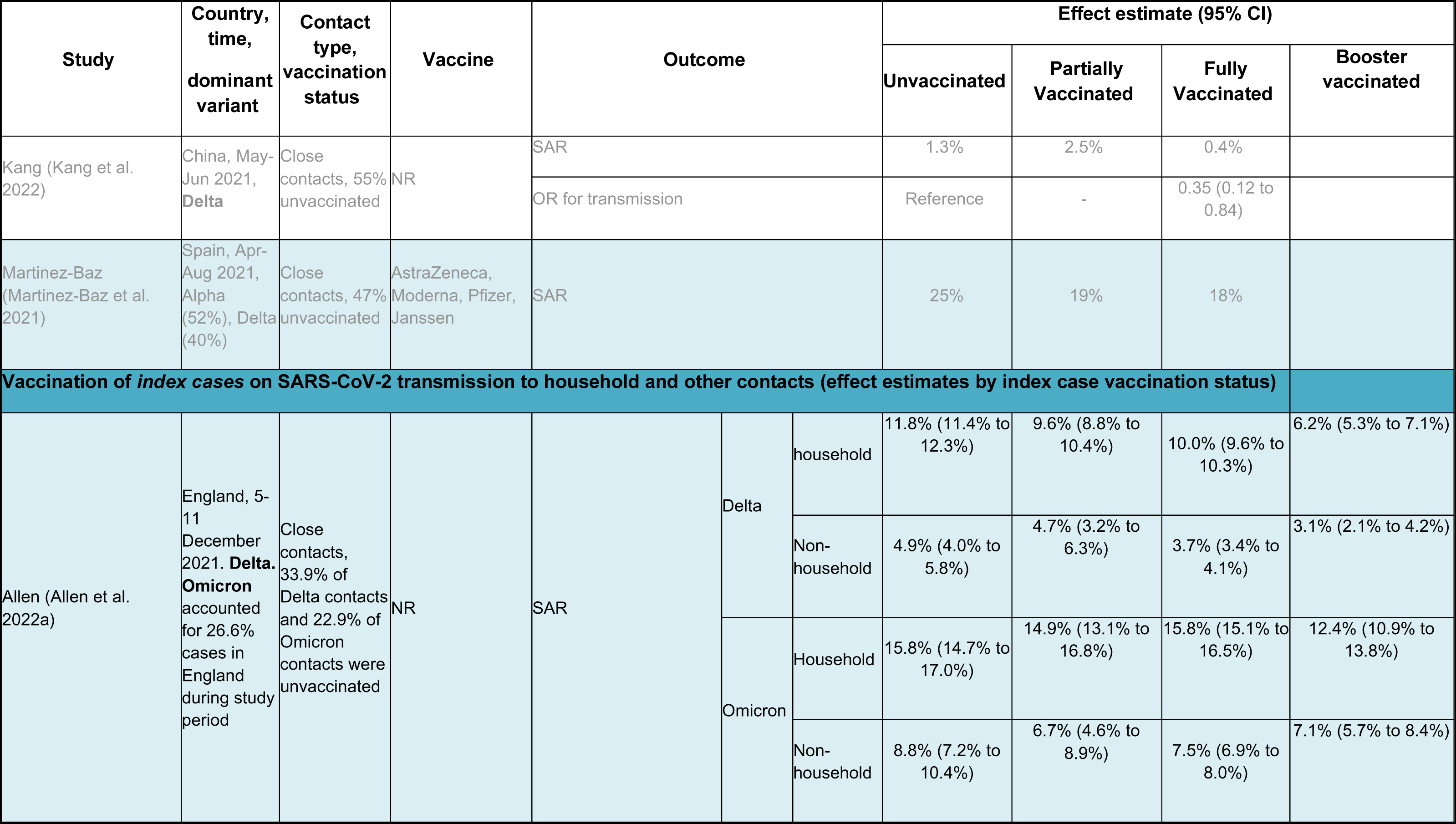
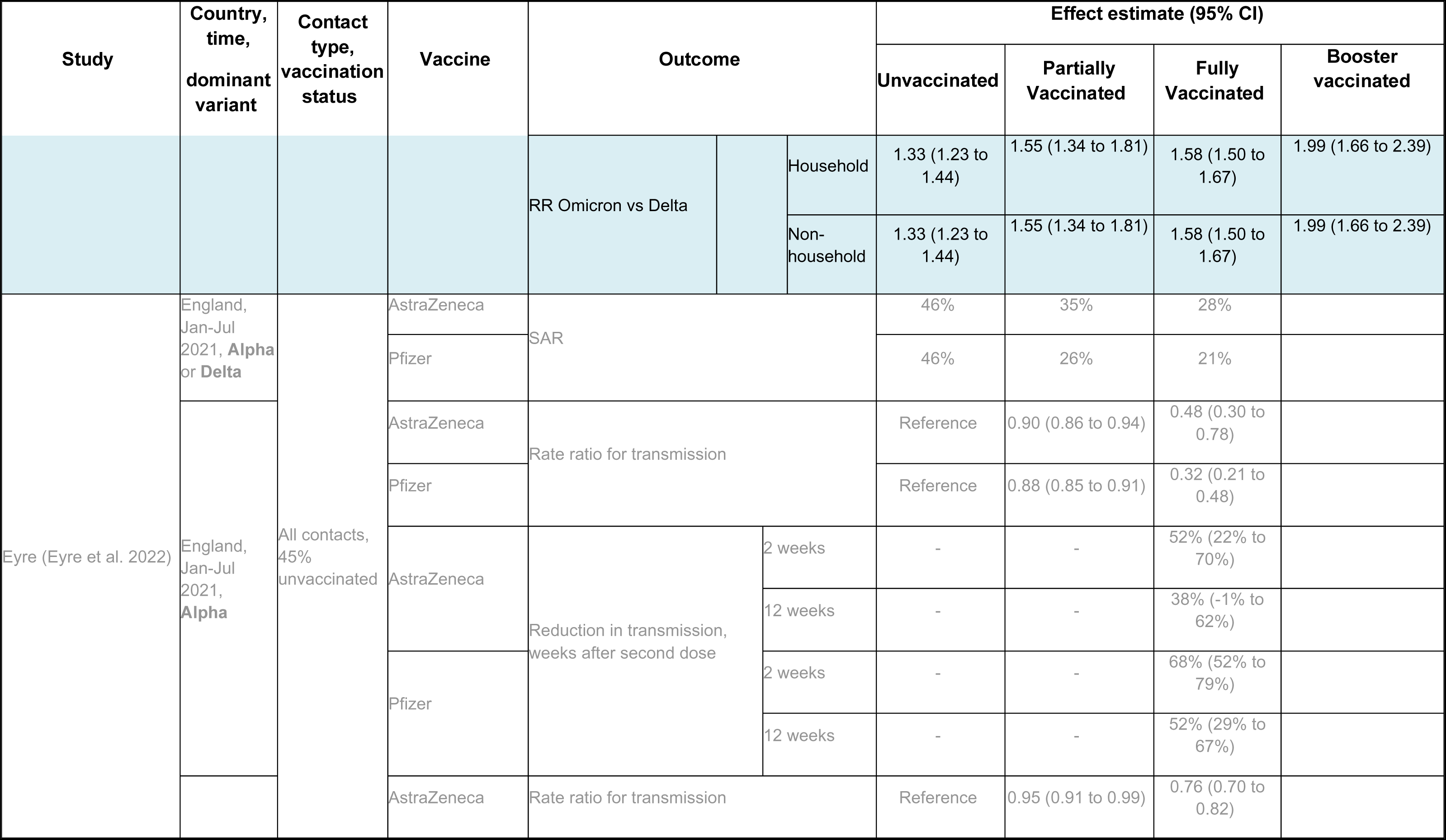
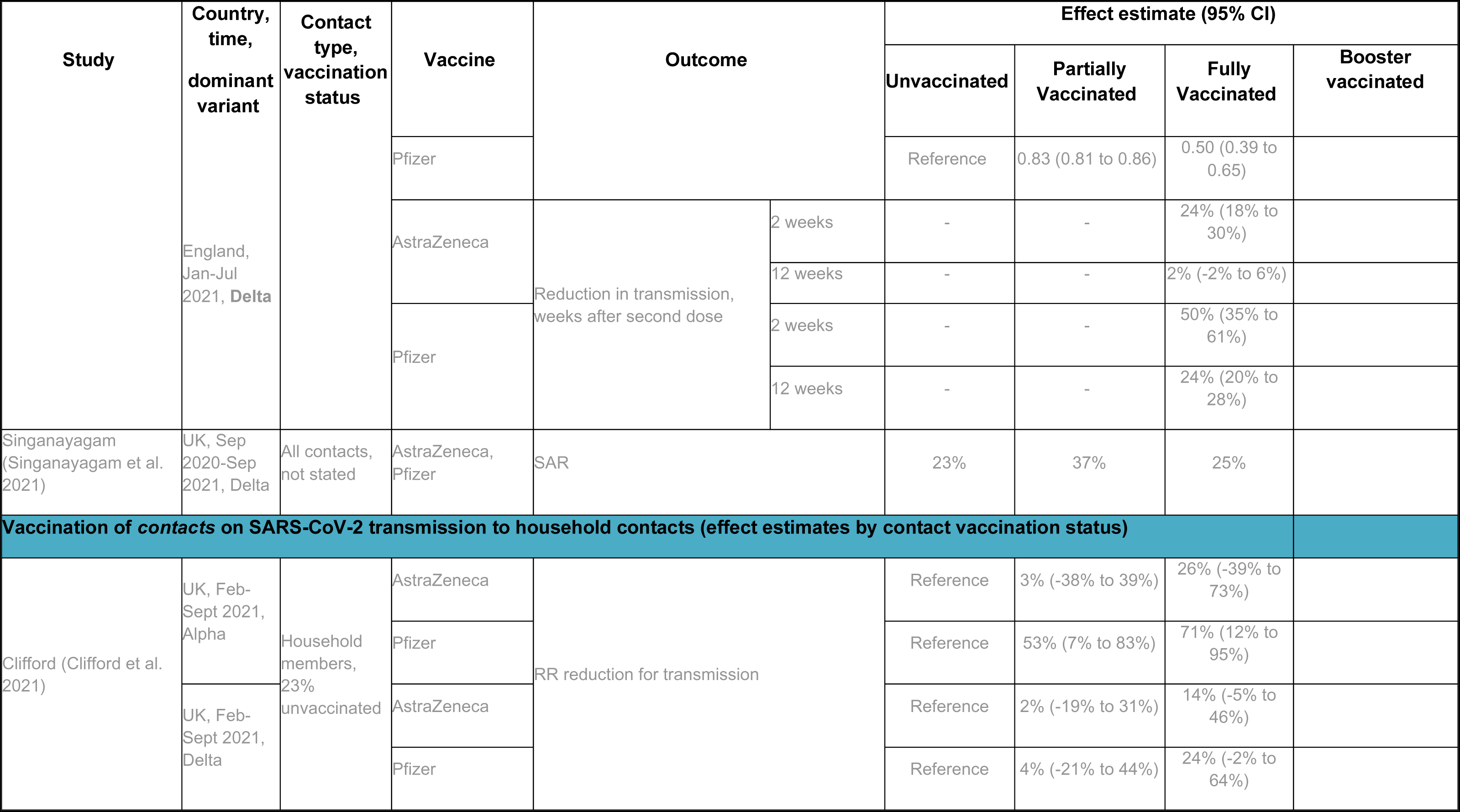
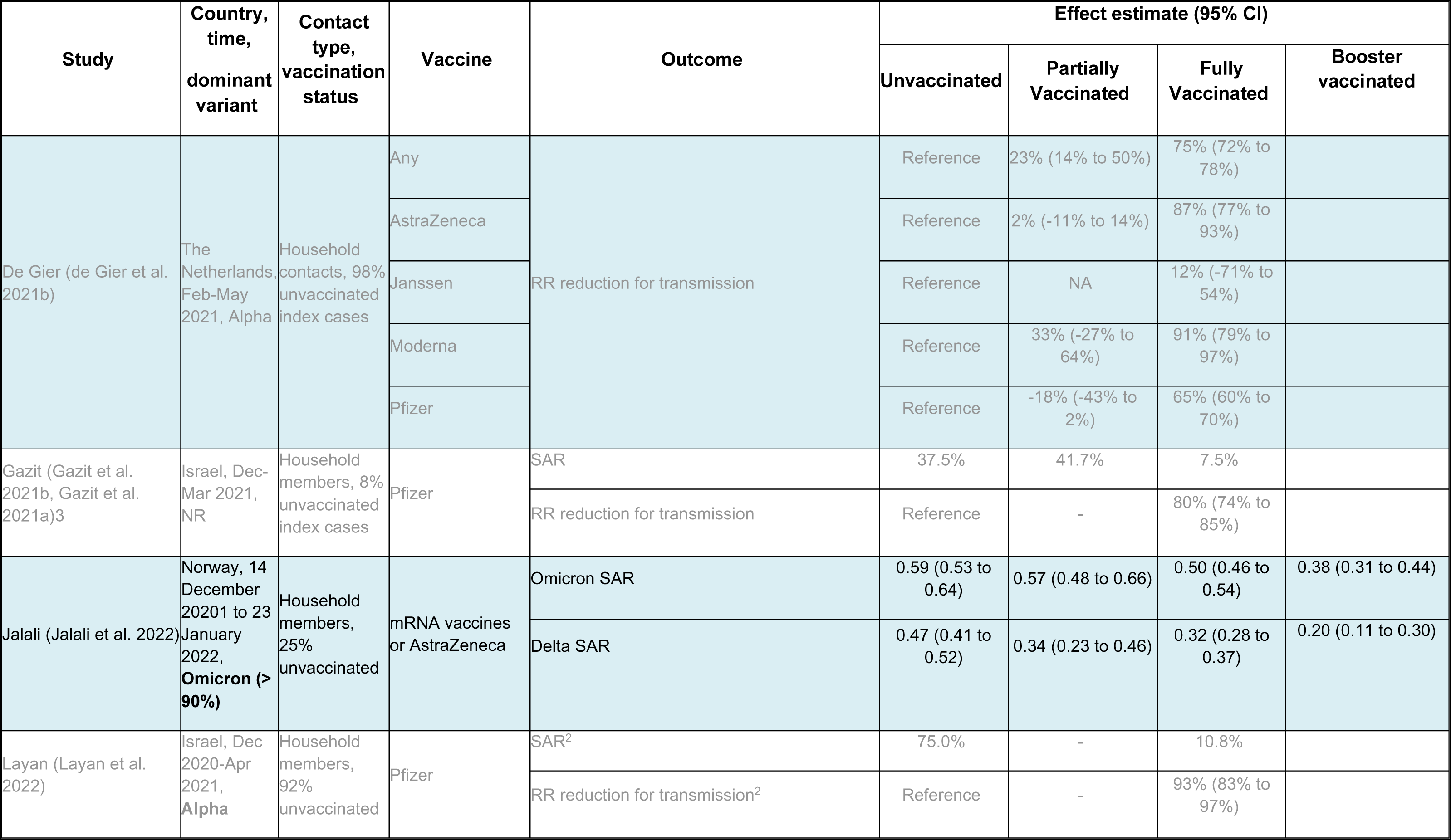
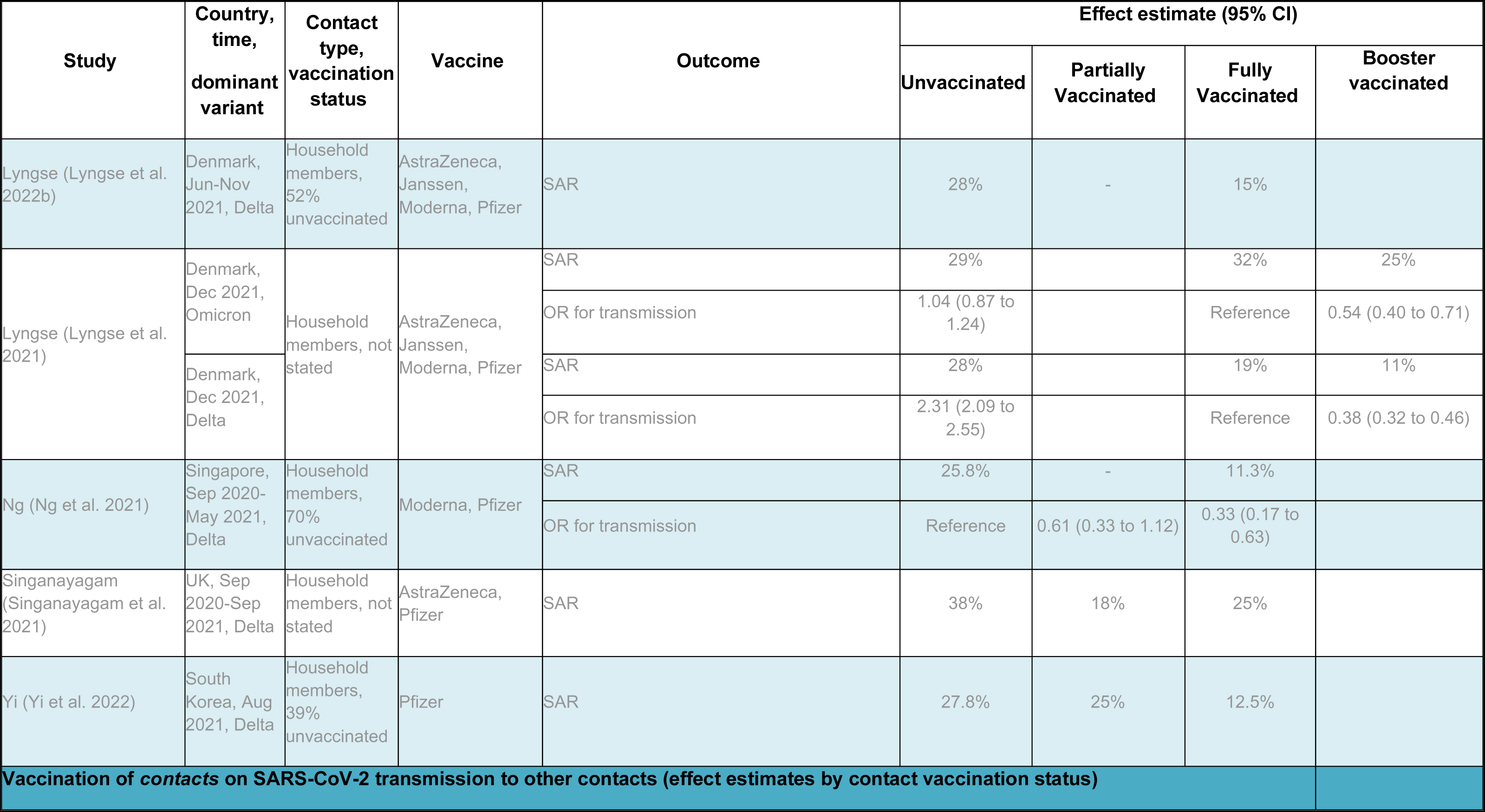
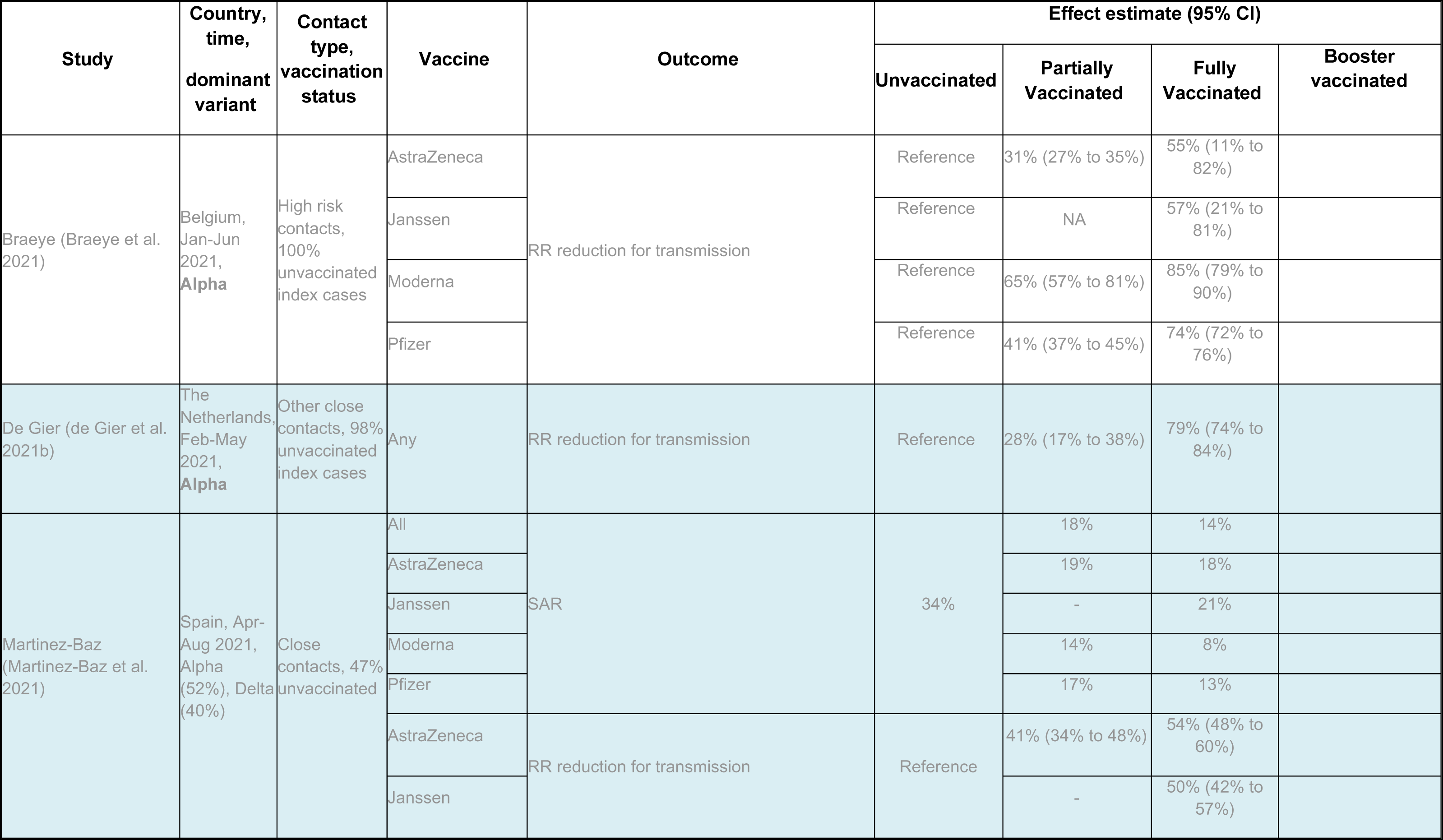
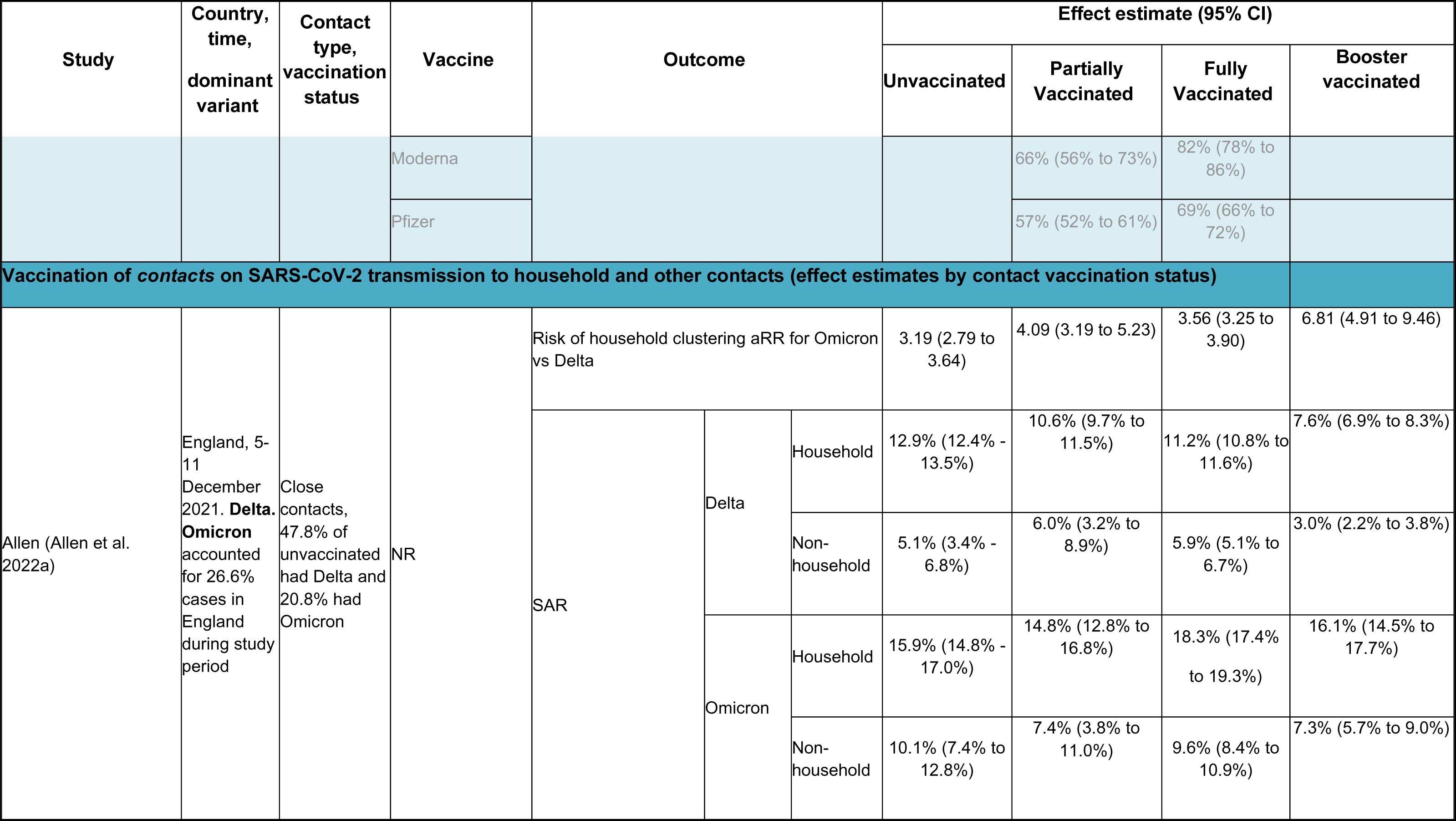
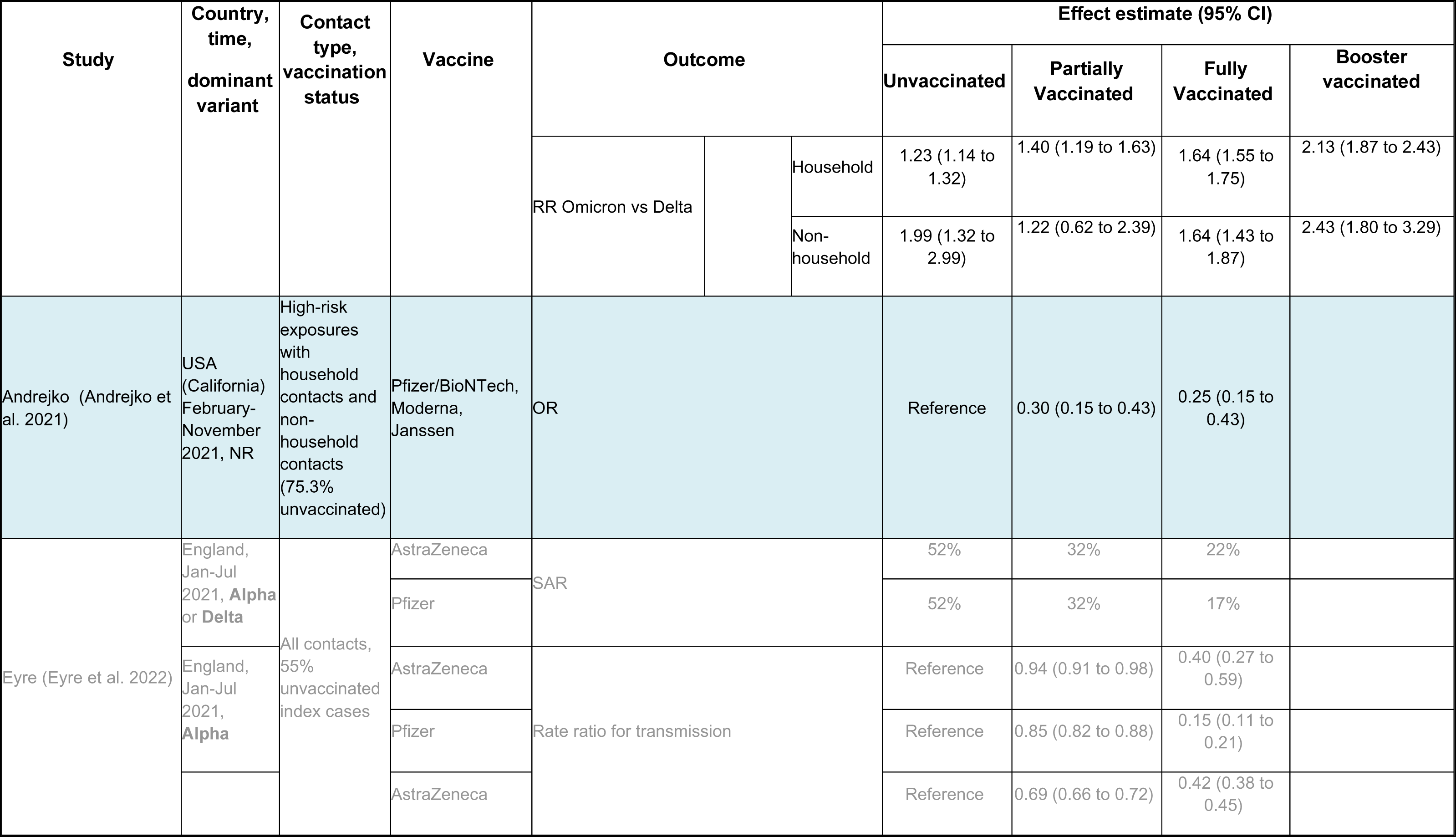
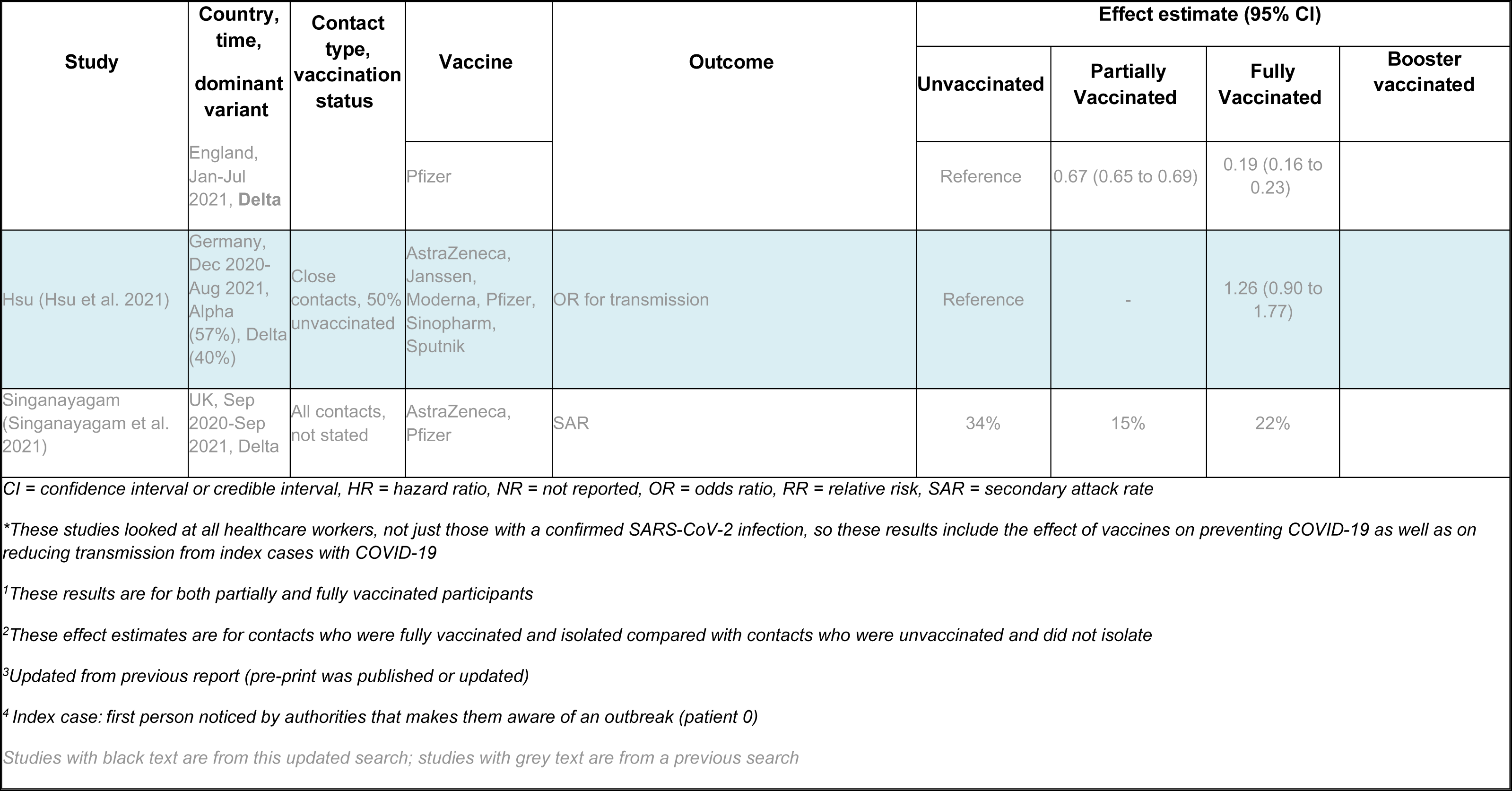
Summary of key findings from transmission studies

**Table 2.**
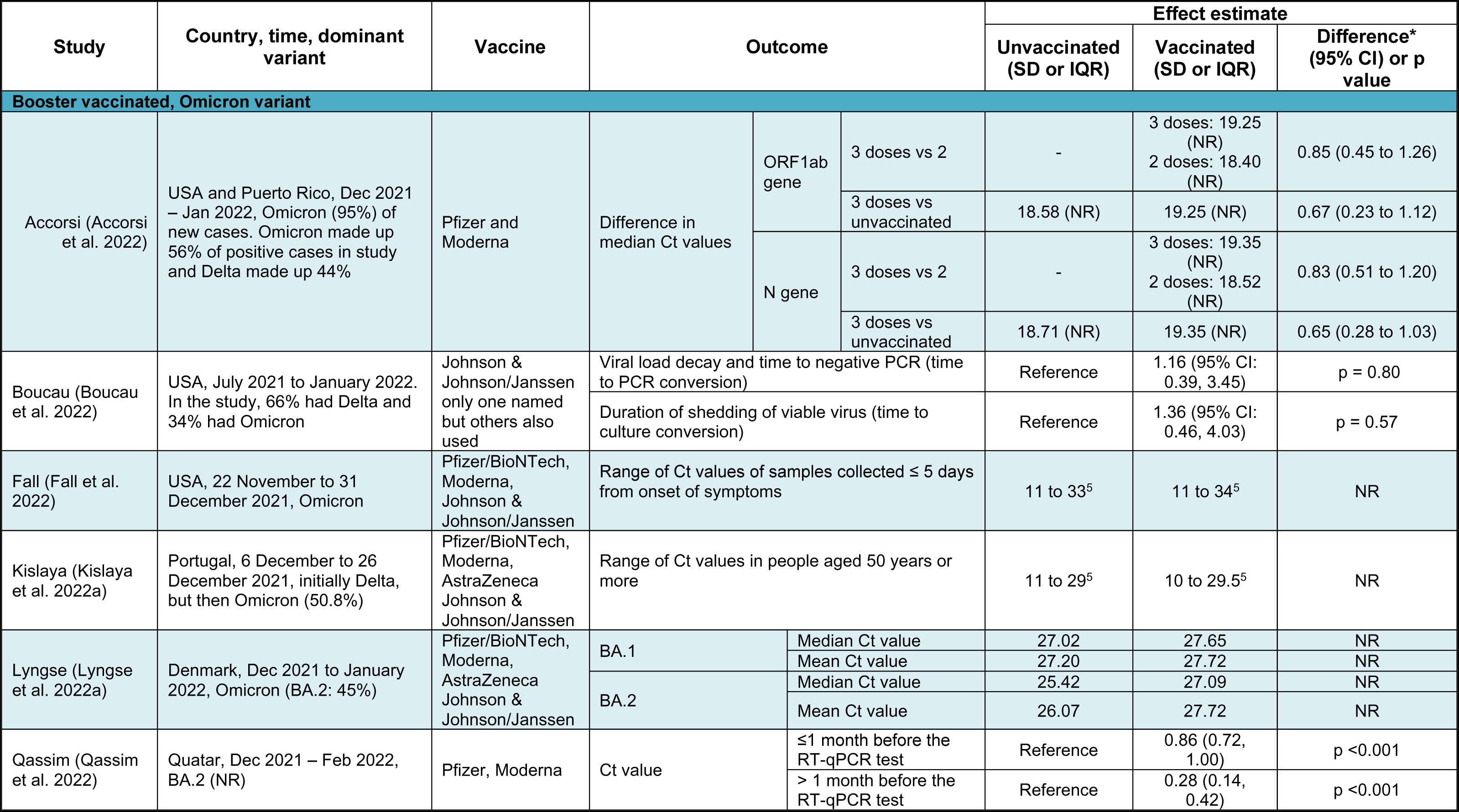
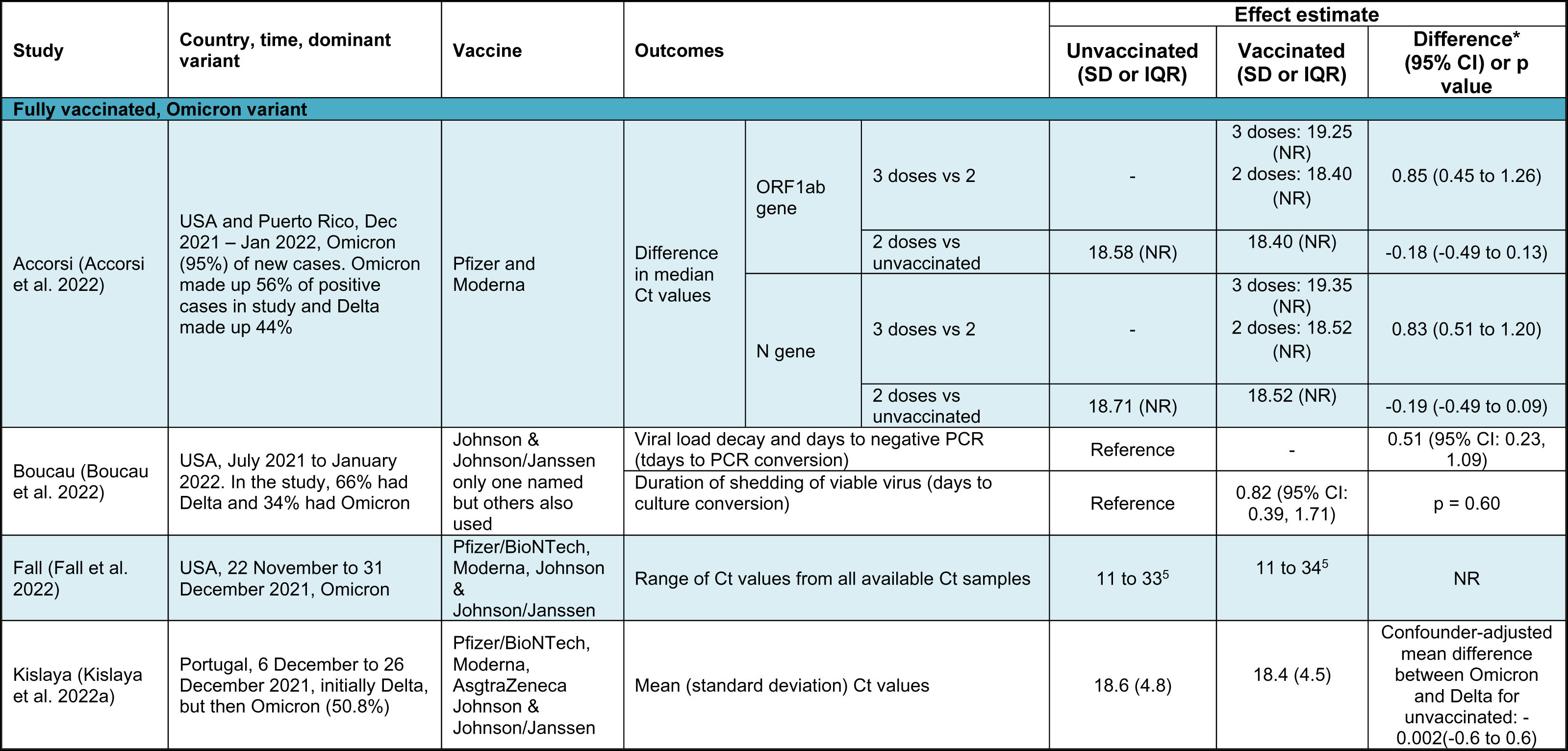
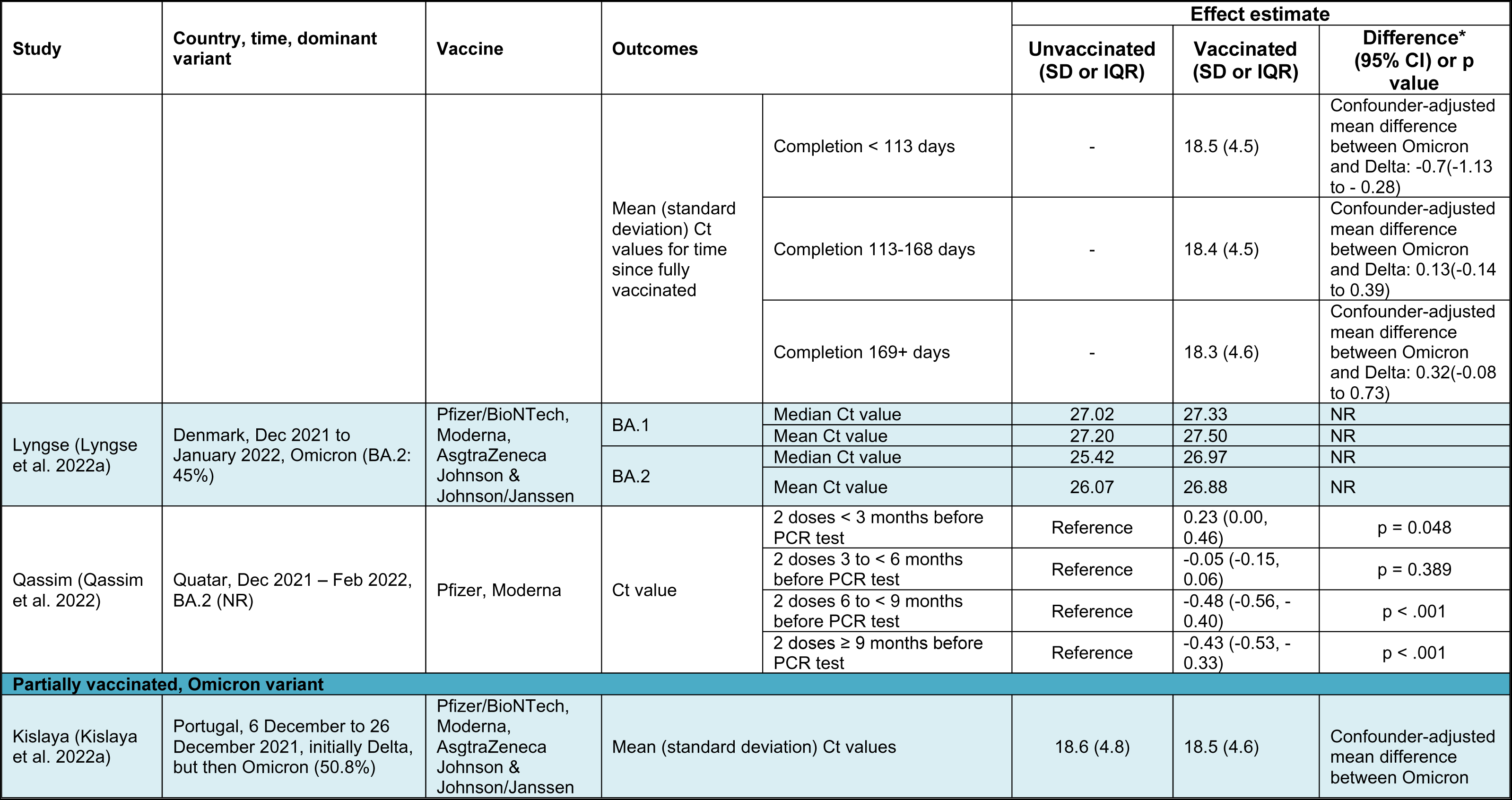
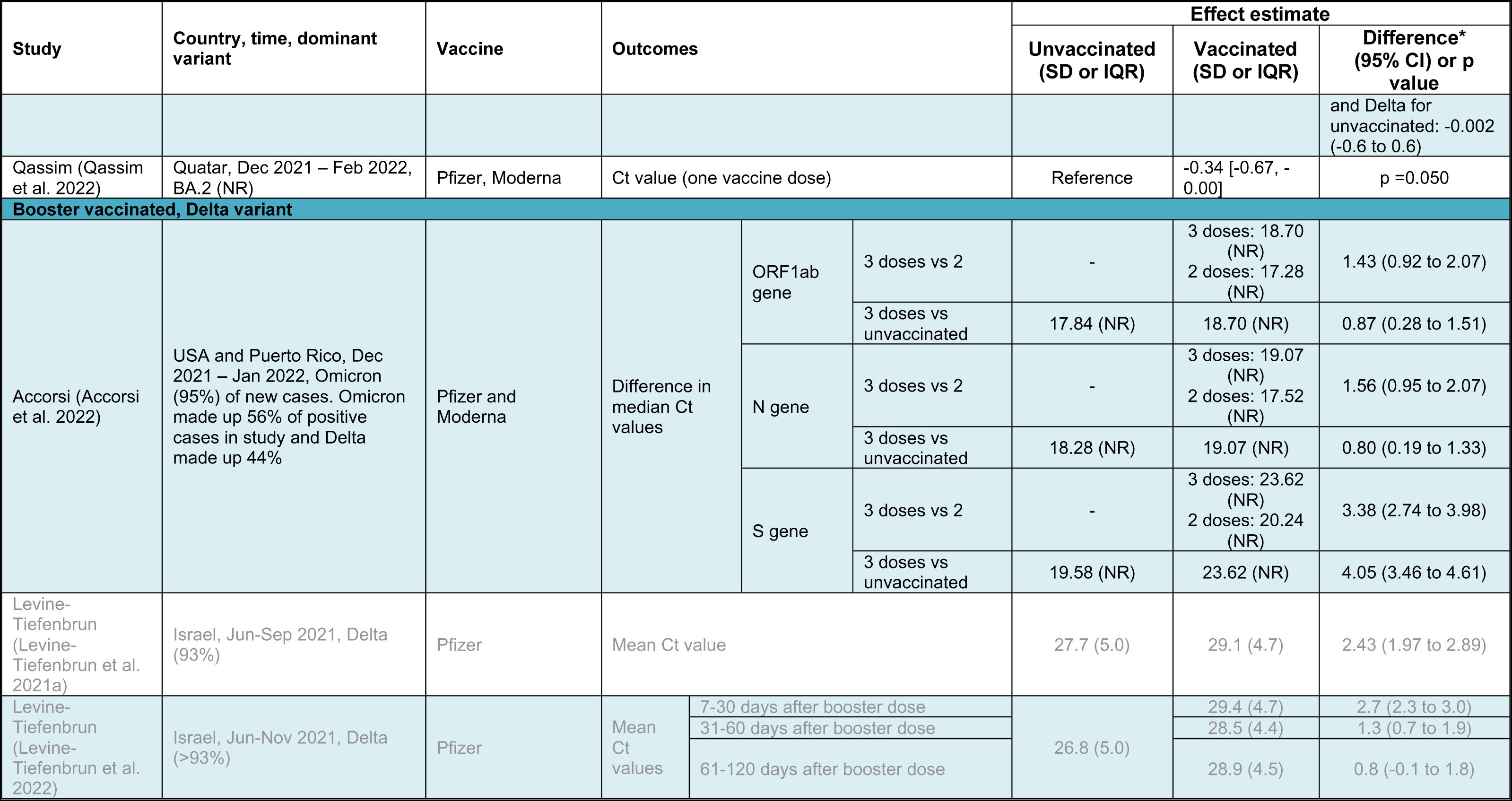
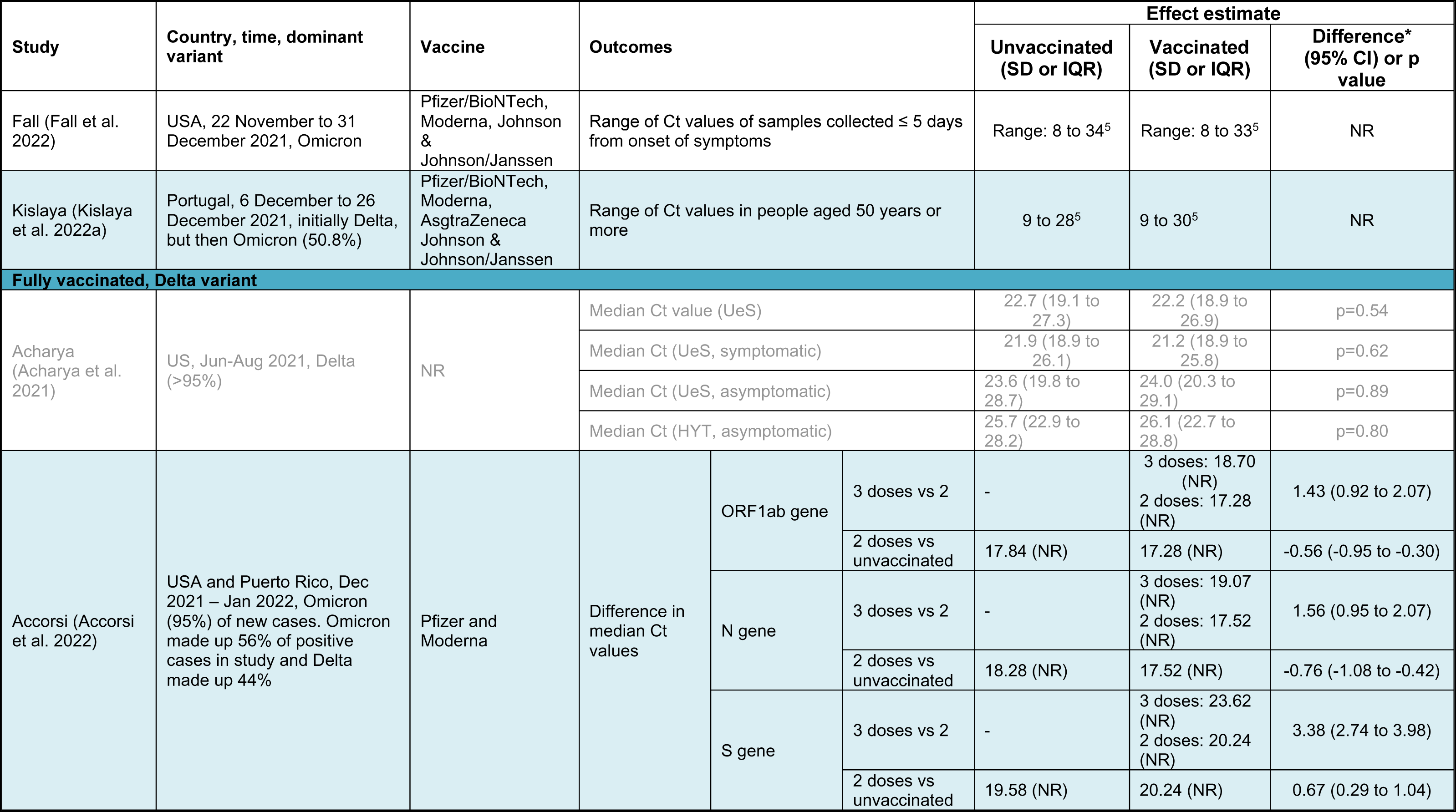
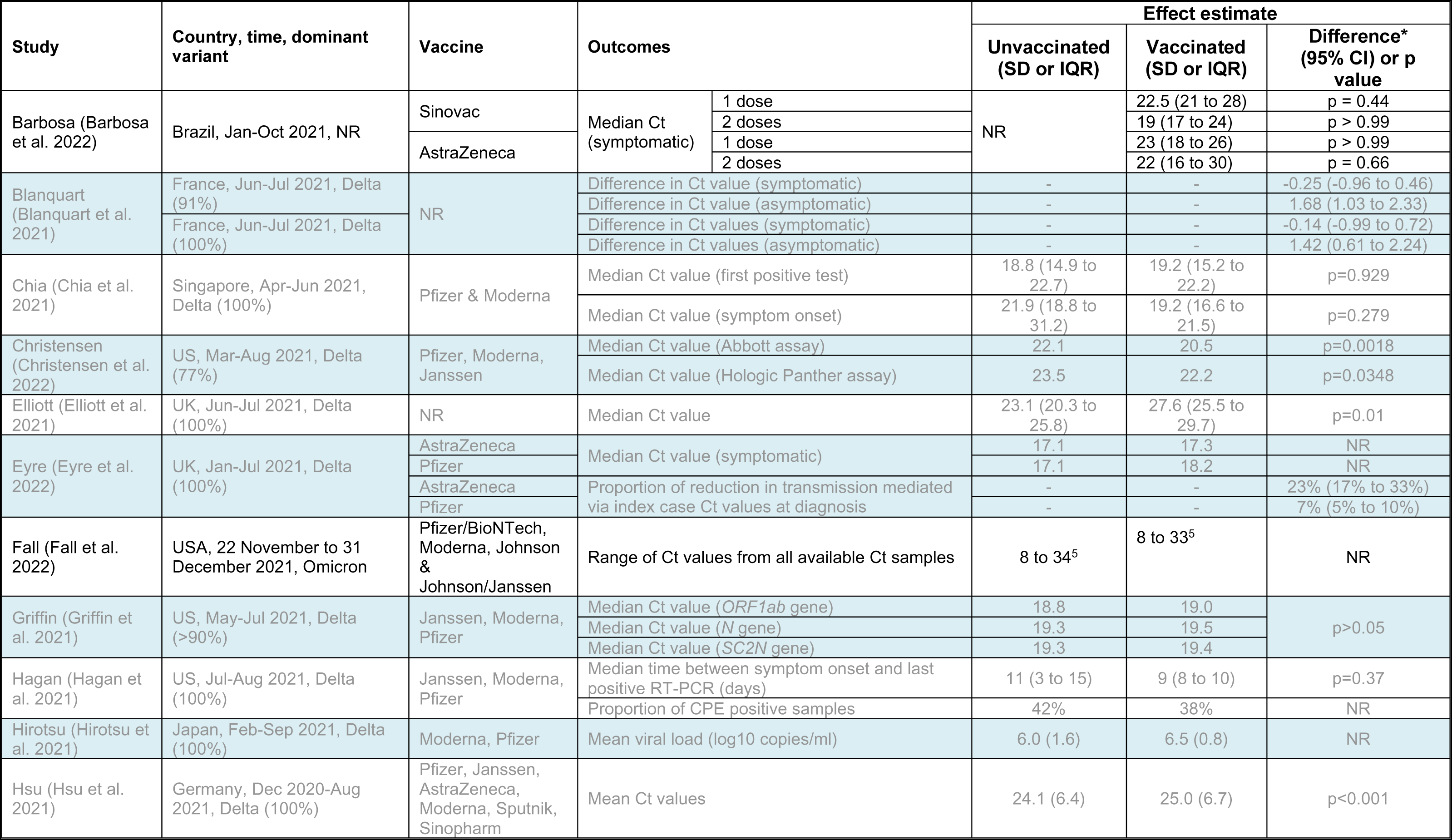
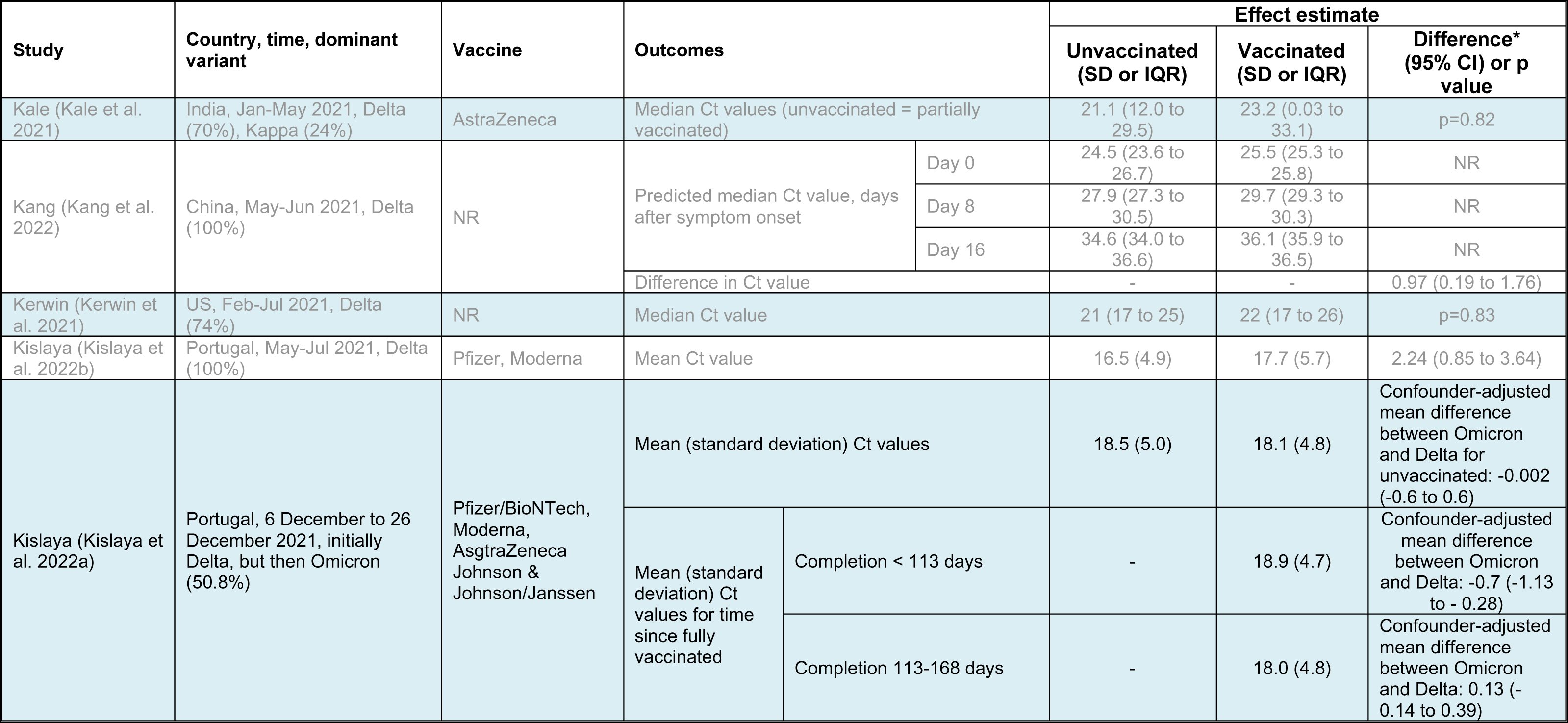
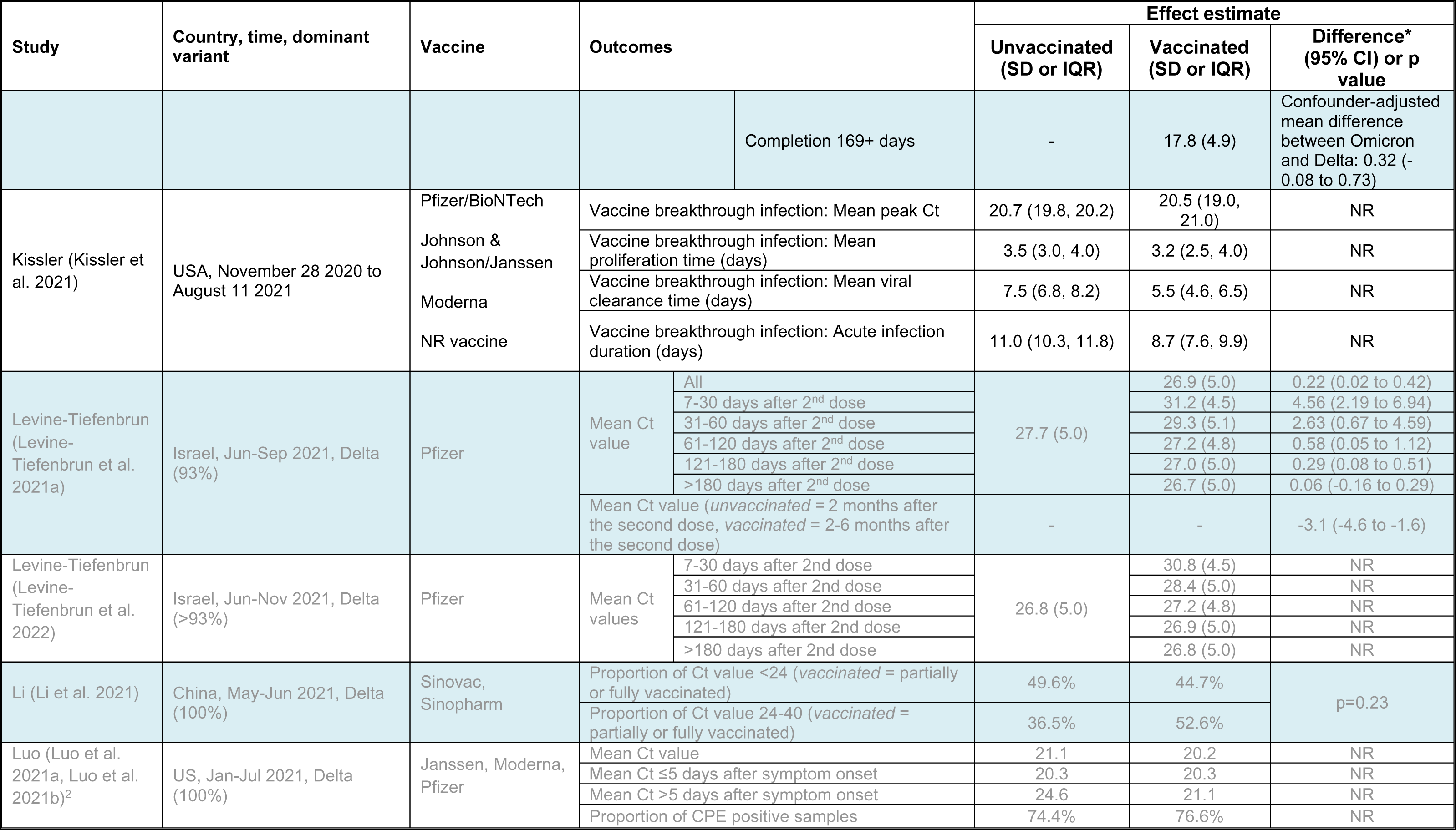
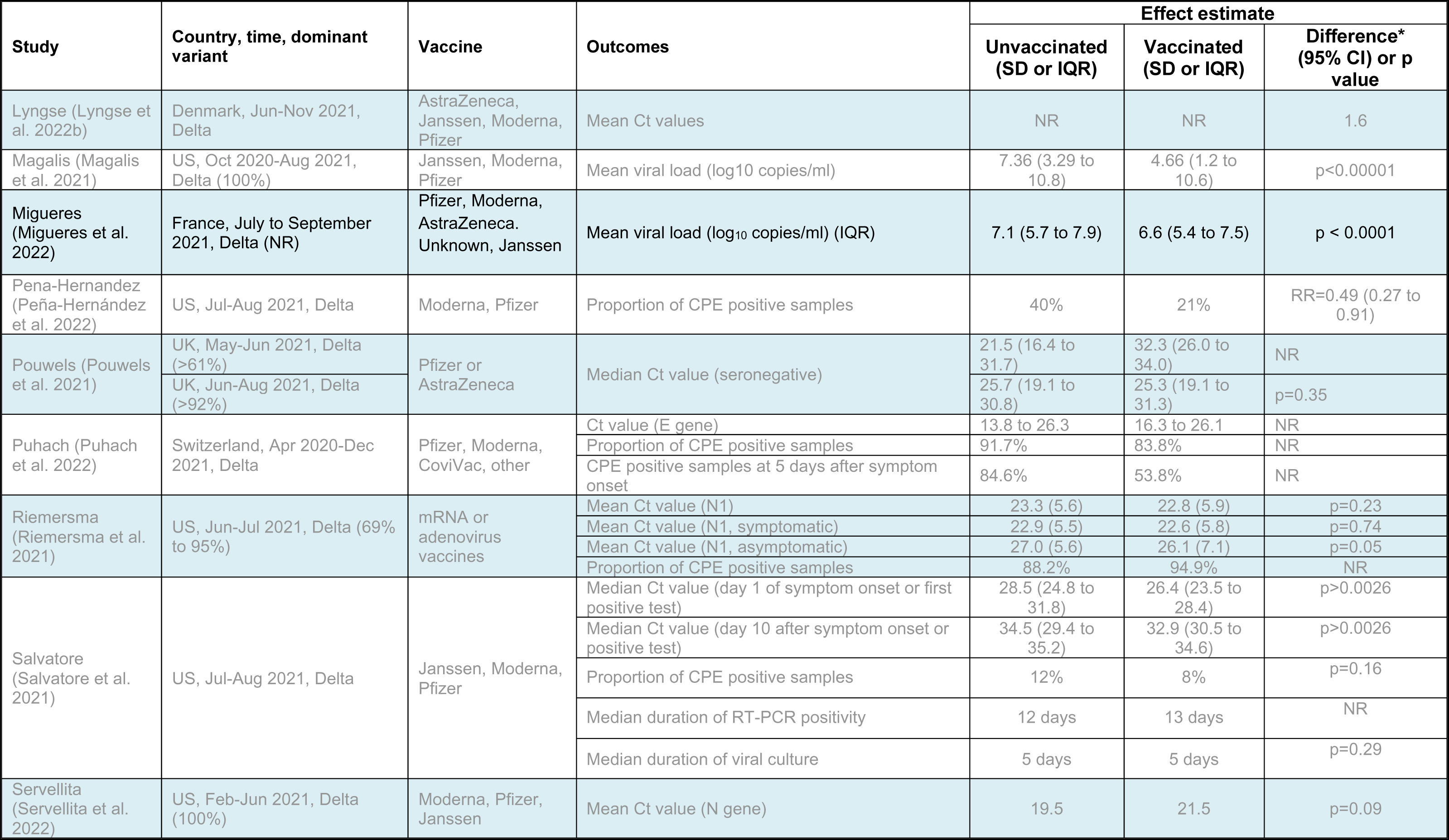
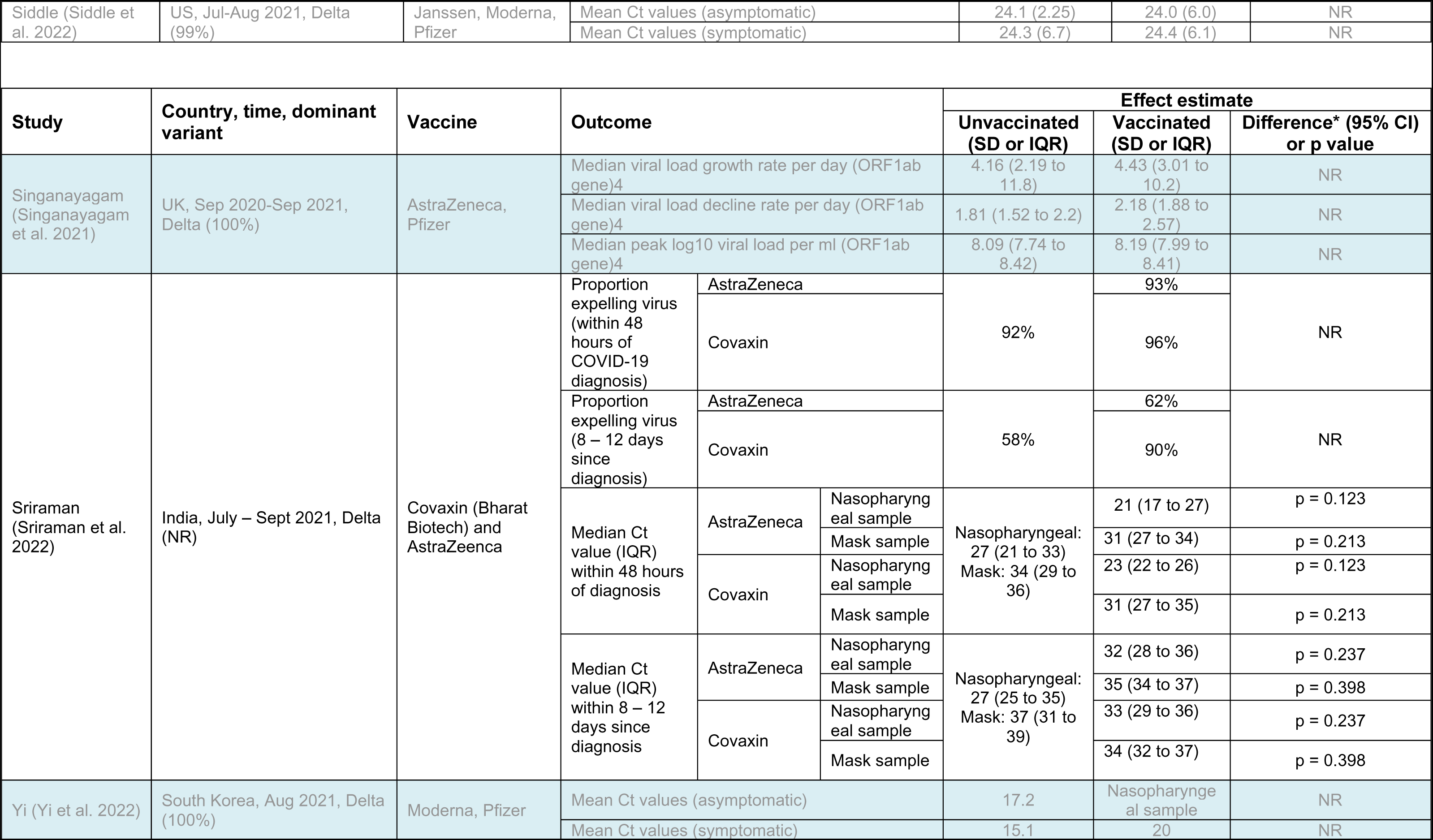
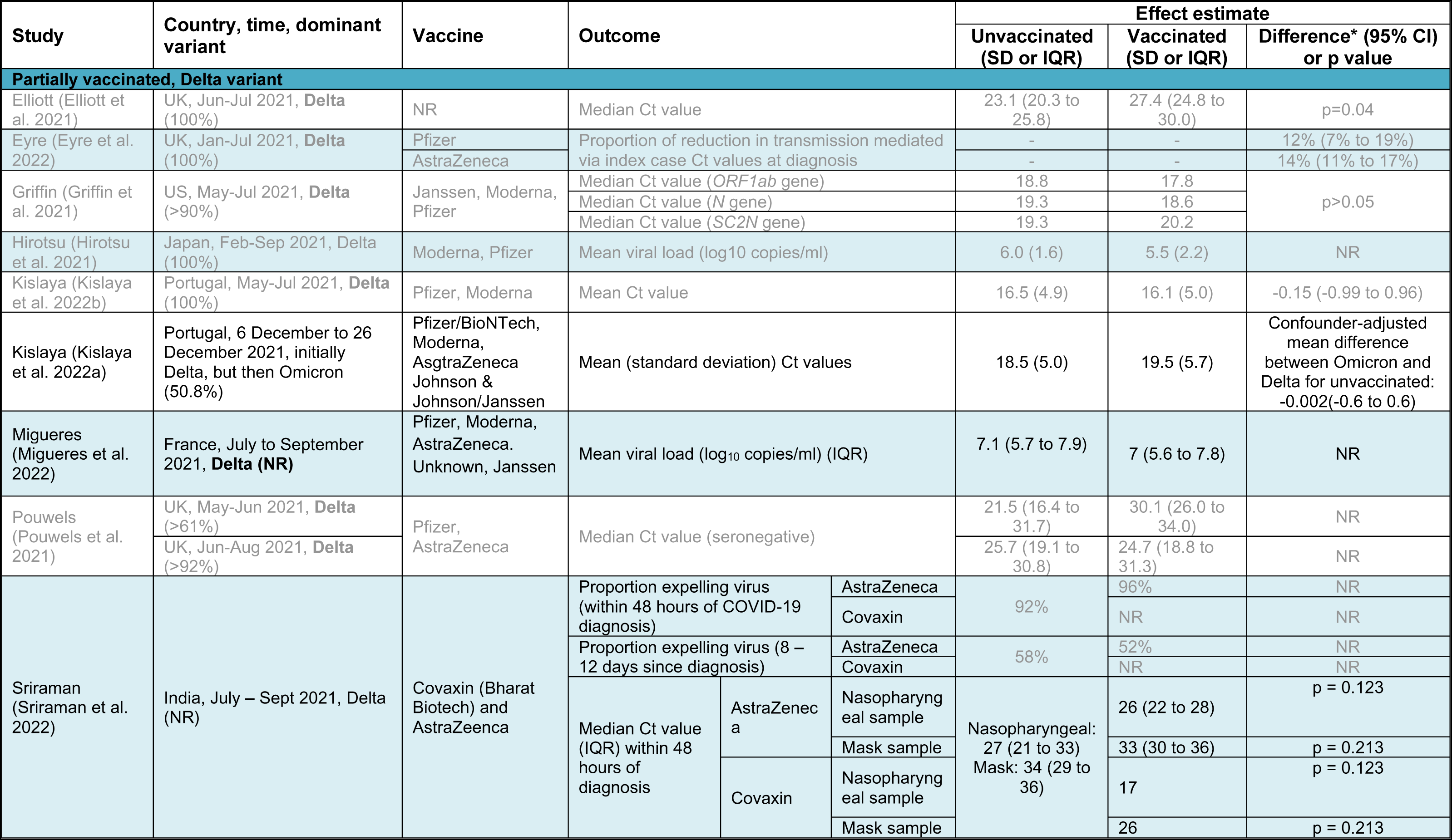
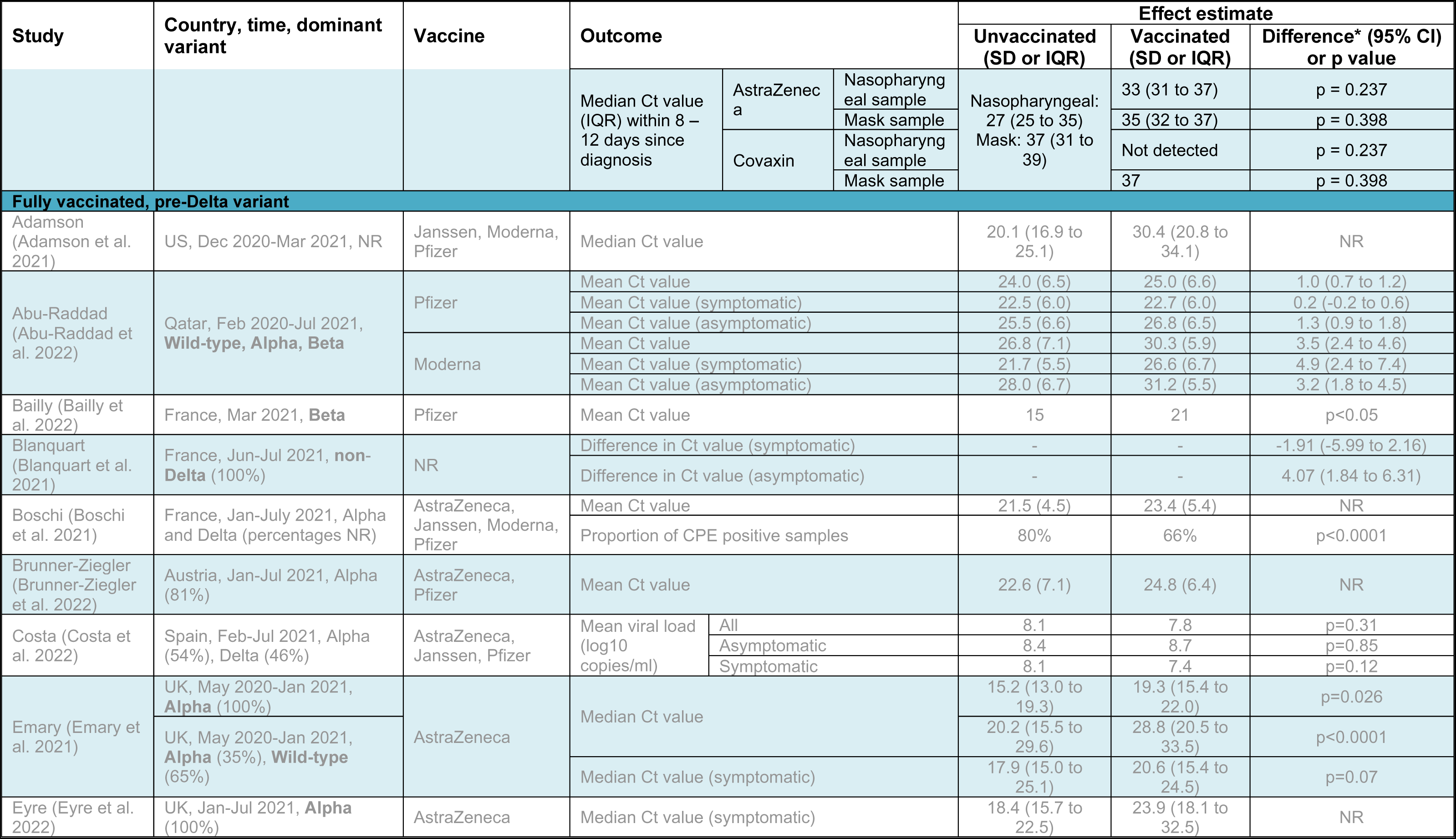
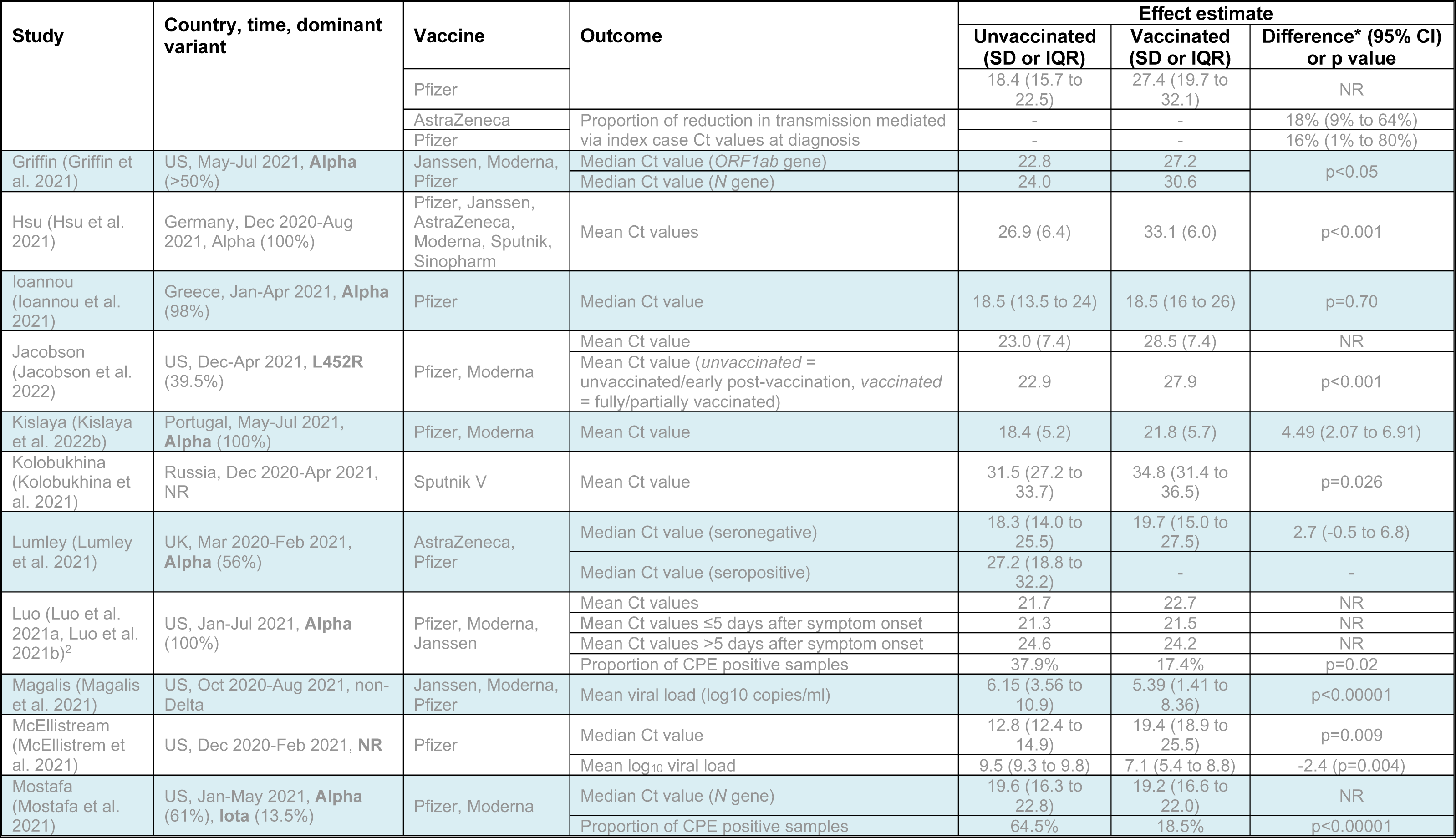
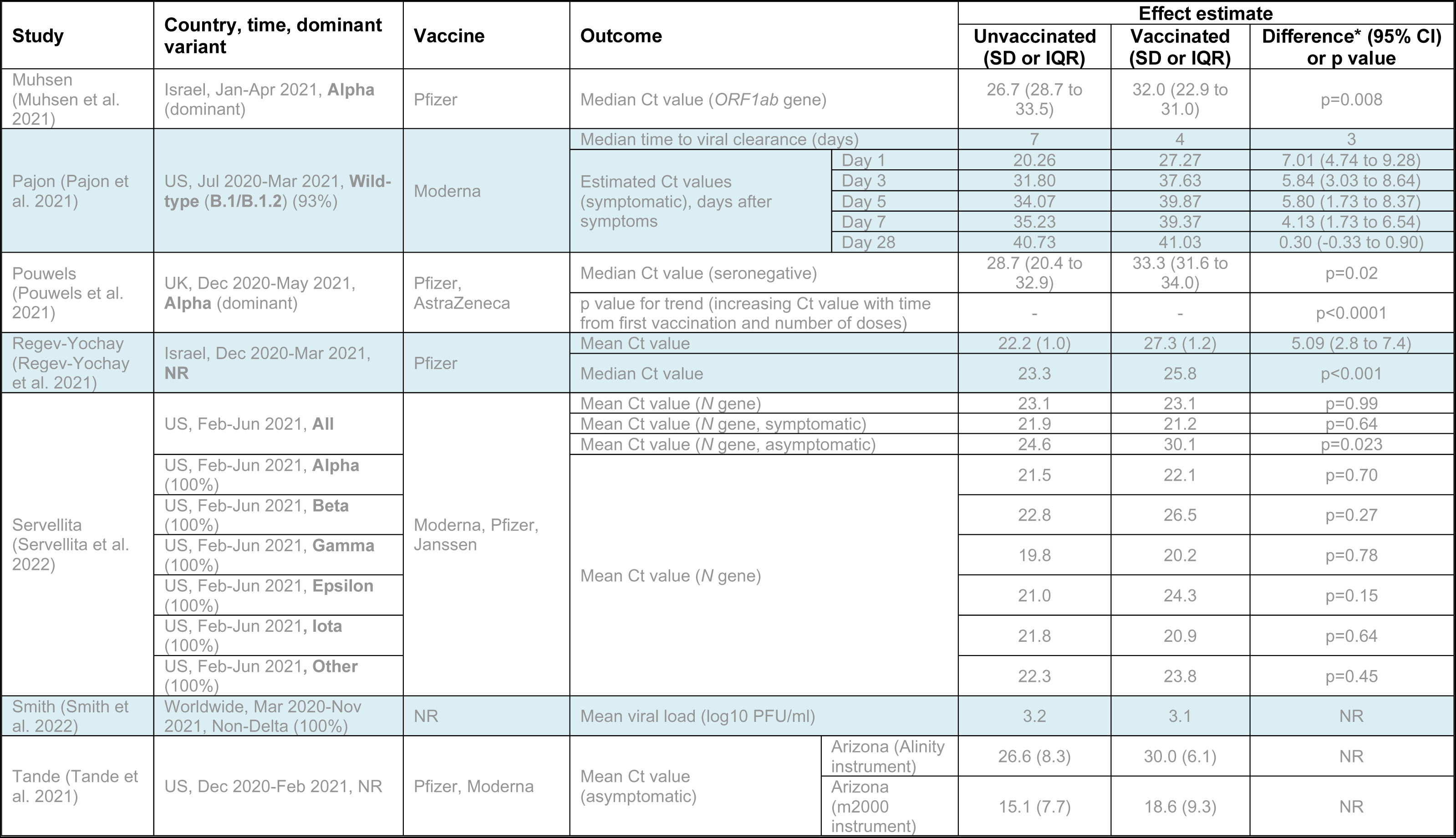
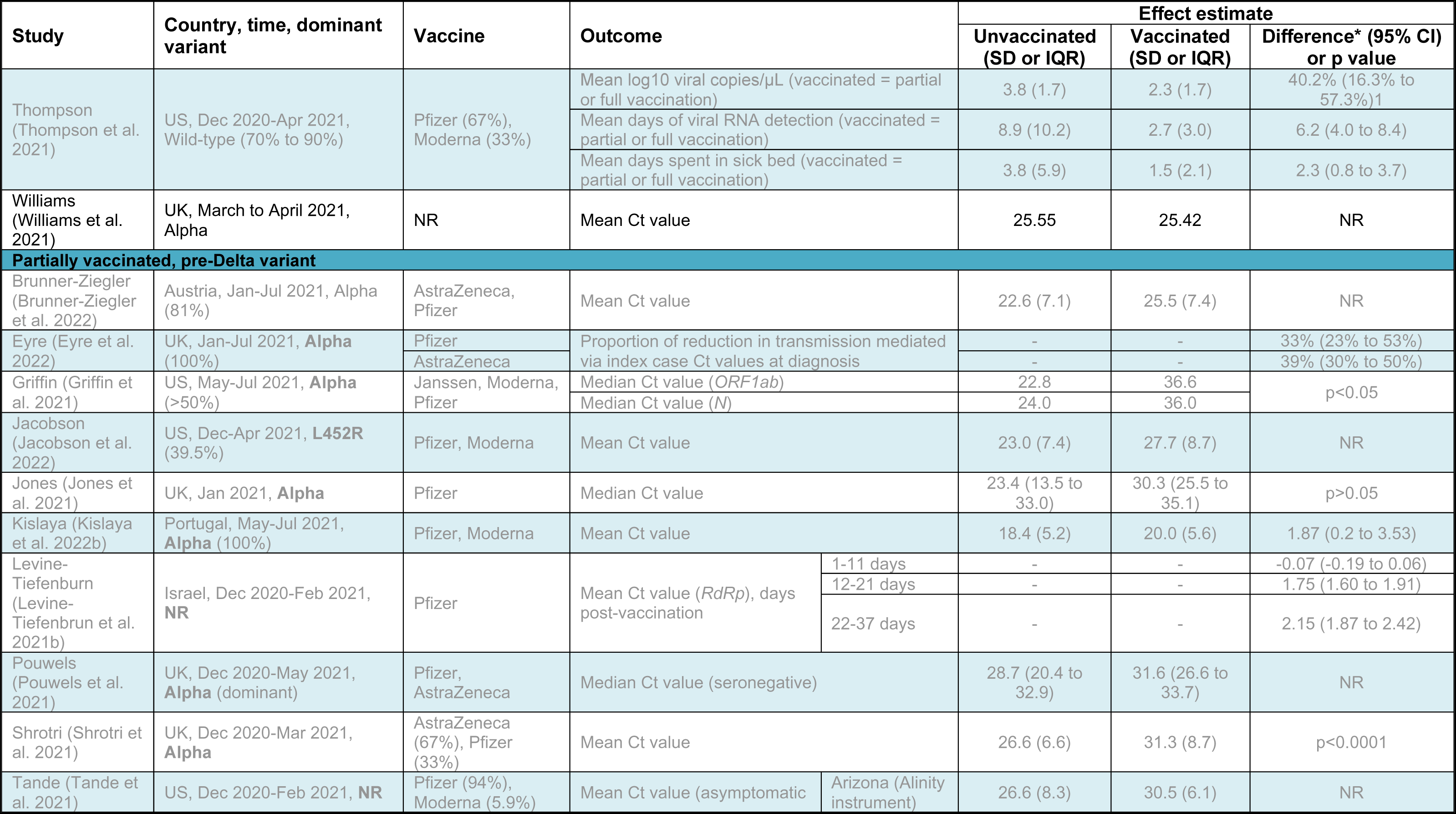
Summary of key findings from studies reporting viral load

## 10. Annex A: Methods

This is an update to a previously published review (UKHSA 2022), and employed a rapid review approach to address the review question:

> Does vaccination against SARS-CoV-2 affect transmission of SARS-CoV-2 to others in the subgroup of people who contract COVID-19 post-vaccination?

We were also interested in the effects of vaccination on transmission according to vaccine type, individual vaccine brands, duration after vaccination, completion of the vaccination course, age and sex of index cases, SARS-CoV-2 variants in index cases, and background SARS-CoV-2 infection rate.

Our rapid review approach follows streamlined systematic methodologies (Tricco et al. 2017). In particular, 10% of the screening on title and abstract were screened in duplicate; full text screening, data extraction and risk of bias assessment were performed by one reviewer and checked by another. The review has been reported according to PRISMA guidelines (Page et al. 2021).

### Protocol

A protocol was produced by the project team before the literature search began, specifying the research question and the inclusion and exclusion criteria. The review was registered prospectively on PROSPERO (CRD42021257125).

### Review questions

1. What is the evidence on SARS-CoV-2 transmission from people who have had one or two doses of a COVID-19 vaccination?
2. How does risk of onward transmission vary with vaccine type, completion of the vaccination course, duration after vaccination, at different baseline community transmission levels and SARS-CoV-2 variant in the vaccinated person?

#### Sources searched

Ovid Medline, Ovid Embase, CENTRAL, medRxiv and SSRN pre-prints, WHO COVID-19 Research Database.

### Search strategy

Searches were conducted for papers published between **12 January 2022** and **15 March 2022**. The previous review included the same search strategy for papers published between 22 October 2021 and 12 January 2022. Studies included in the previous review are also included in this review.

Search terms covered key aspects of the review question. The search strategy for Ovid Medline is presented in **Box A.1**. Additionally, we checked reference lists of relevant systematic reviews and evidence summaries and consulted with topic experts. All that had been identified as pre-prints as of **12 January 2022** were last checked and updated (if necessary) on 6 April 2022.

#### Box A.1.

##### Search strategy Ovid Medline

1. vaccinat*.tw,kw.
2. vaccine*.tw,kw.
3. previously-vaccin*.tw,kw.
4. post-vaccin*.tw,kw.
5. early-vaccin*.tw,kw.
6. late-vaccin*.tw,kw.
7. moderna.tw,kw.
8. mRNA-1273.tw,kw.
9. pfizer.tw,kw.
10. BNT162b2.tw,kw.
11. JNJ-78436735.tw,kw.
12. "Johnson & Johnson*".tw,kw.
13. Astrazeneca.tw,kw.
14. Oxford-Astrazeneca.tw,kw.
15. AZD 1222.tw,kw.
16. AZD1222.tw,kw.
17. BNT 162b2.tw,kw.
18. ChAdOx1.tw,kw.
19. Novavax.tw,kw.
20. NVX-CoV2373.tw,kw.
21. Sputnik V.tw,kw.
22. Ad26.tw,kw.
23. "Ad26.COV2".tw,kw.
24. Ad5.tw,kw.
25. Janssen.tw,kw.
26. Sinovac.tw,kw.
27. sinopharm.tw,kw.
28. covaxin.tw,kw.
29. exp Vaccination/
30. COVID-19 Vaccines/
31. 1 or 2 or 3 or 4 or 5 or 6 or 7 or 8 or 9 or 10 or 11 or 12 or 13 or 14 or 15 or 16 or 17 or 18 or 19 or 20 or 21 or 22 or 23 or 24 or 25 or 26 or 27 or 28 or 29 or 30
32. (breakthrough or break through).tw,kw.
33. transmiss*.tw,kw.
34. transmit*.tw,kw.
35. viral load*.tw,kw.
36. viral burden.tw,kw.
37. ((severity or severe) adj2 (disease or illness)).tw,kw.
38. Viral Load/
39. exp Disease Transmission, Infectious/
40. 32 or 33 or 34 or 35 or 36 or 37 or 38 or 39
41. exp coronavirus/
42. exp Coronavirus Infections/
43. COVID-19/
44. ((corona* or corono*) adj1 (virus* or viral* or virinae*)).ti,ab,kw.
45. (coronavirus* or coronovirus* or coronavirinae* or CoV or HCoV*).ti,ab,kw.
46. covid*.nm.
47. (2019-nCoV or 2019nCoV or nCoV2019 or nCoV-2019 or COVID-19 or COVID19 or CORVID-19 or CORVID19 or WN-CoV or WNCoV or HCoV-19 or HCoV19 or 2019 novel* or Ncov or n-cov or SARS-CoV-2 or SARSCoV-2 or SARSCoV2 or SARS-CoV2 or SARSCov19 or SARS-Cov19 or SARSCov-19 or SARS-Cov-19 or Ncovor or Ncorona* or Ncorono* or NcovWuhan* or NcovHubei* or NcovChina* or NcovChinese* or SARS2 or SARS-2 or SARScoronavirus2 or SARS-coronavirus-2 or SARScoronavirus 2 or SARS coronavirus2 or SARScoronovirus2 or SARS-coronovirus-2 or SARScoronovirus 2 or SARS coronovirus2).ti,ab,kw.
48. (respiratory* adj2 (symptom* or disease* or illness* or condition*) adj10 (Wuhan* or Hubei* or China* or Chinese* or Huanan*)).ti,ab,kw.
49. ((seafood market* or food market* or pneumonia*) adj10 (Wuhan* or Hubei* or China* or Chinese* or Huanan*)).ti,ab,kw.
50. ((outbreak* or wildlife* or pandemic* or epidemic*) adj1 (Wuhan* or Hubei or China* or Chinese* or Huanan*)).ti,ab,kw.
51. or/41-50
52. 31 and 40 and 51
53. COVID-19/tm [Transmission]
54. 31 and 53
55. COVID-19 Vaccines/
56. 40 and 5557.
57. COVID-19/vi [Virology]
58. 31 and 57
59. 52 or 54 or 56 or 58

### Inclusion and exclusion criteria

Article eligibility criteria are summarised in **Table A.1.**

In the protocol, we stated we would include disease severity as an outcome. However, as more transmission evidence became available, the need to include disease severity as a secondary outcome became less necessary, and as with the previous review, we focussed this review on transmission and viral load only. We also stated in the protocol that we would exclude studies where the only index cases were children, as they were not eligible for vaccination when the protocol was written. As in the previous review, we have removed this exclusion criteria. We have also removed the need for contacts to be unvaccinated in transmission studies from the inclusion criteria.

**Table A. 1.**
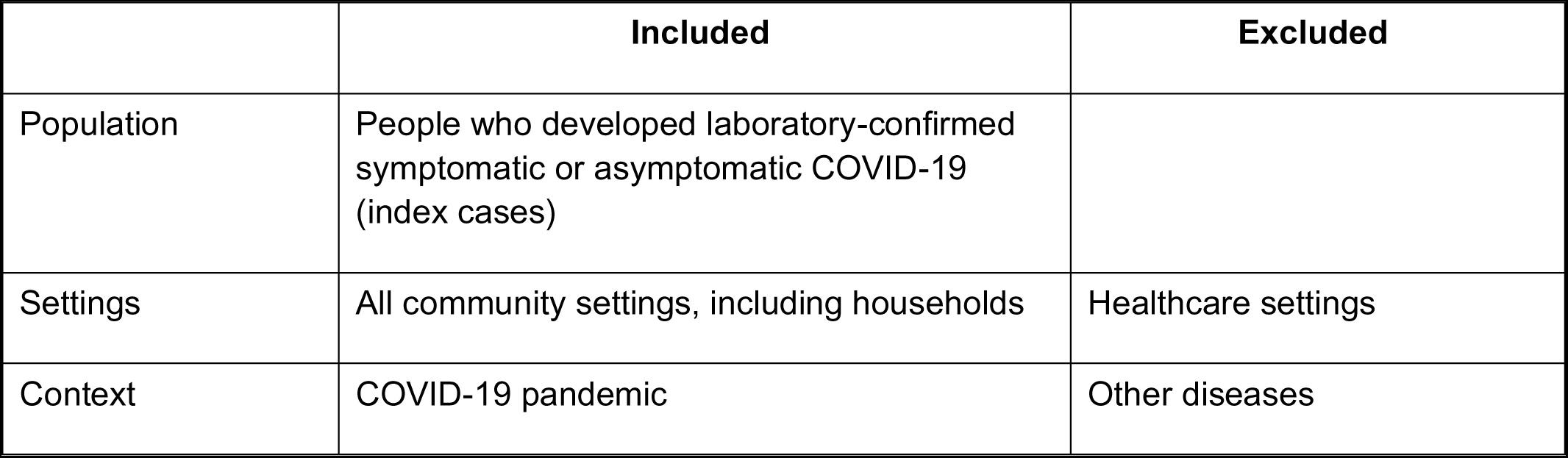
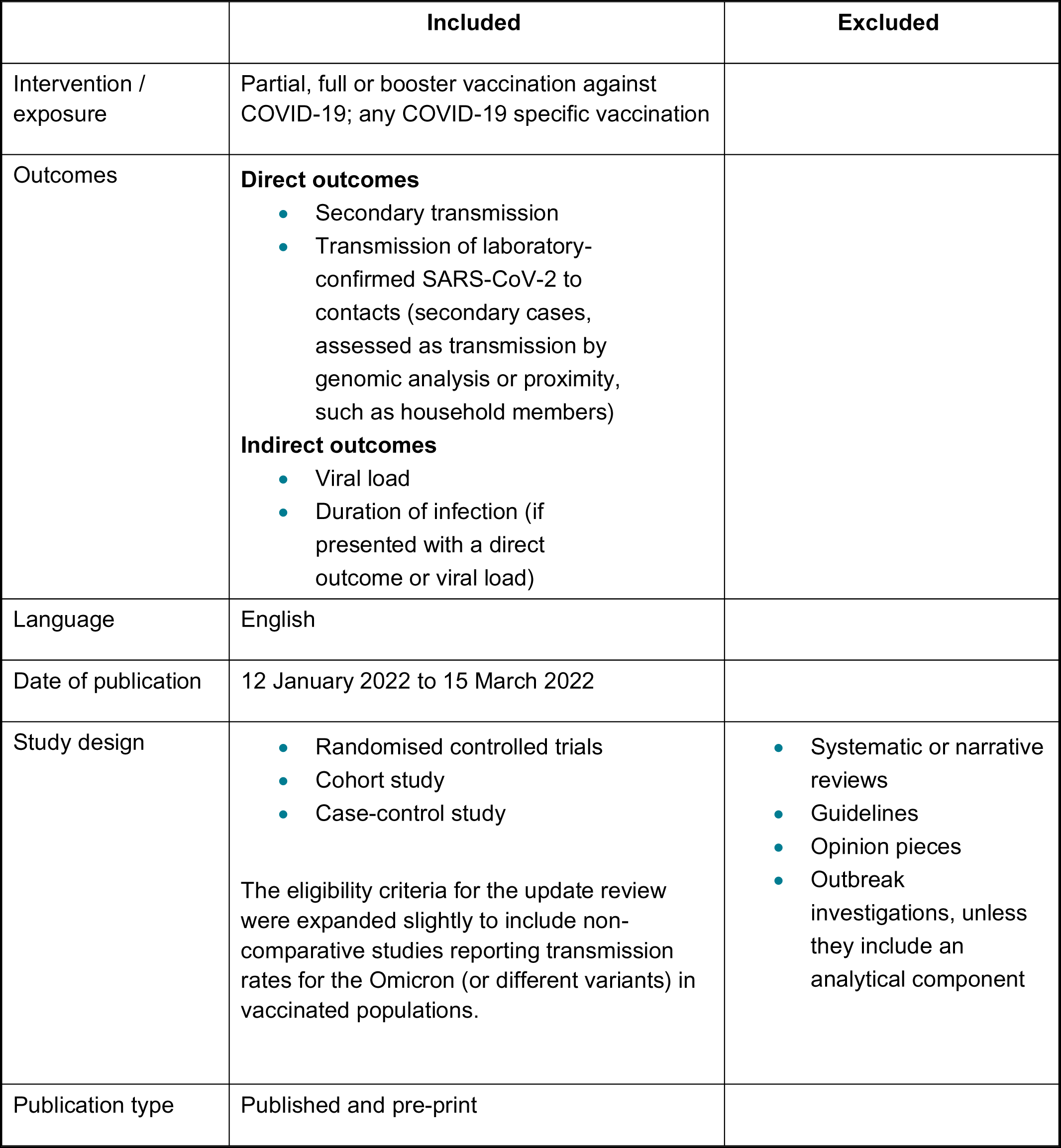
Inclusion and exclusion criteria Screening

### Screening

Title and abstract screening were completed by 2 reviewers: 10% of the eligible studies were screened in duplicate (disagreements were resolved by discussion) and the remainder were screened by 1 reviewer.

Full text screening was completed by one reviewer and checked by a second. The PRISMA diagram showing the flow of citations is provided in **Figure A.1**.

**Figure A. 1.**
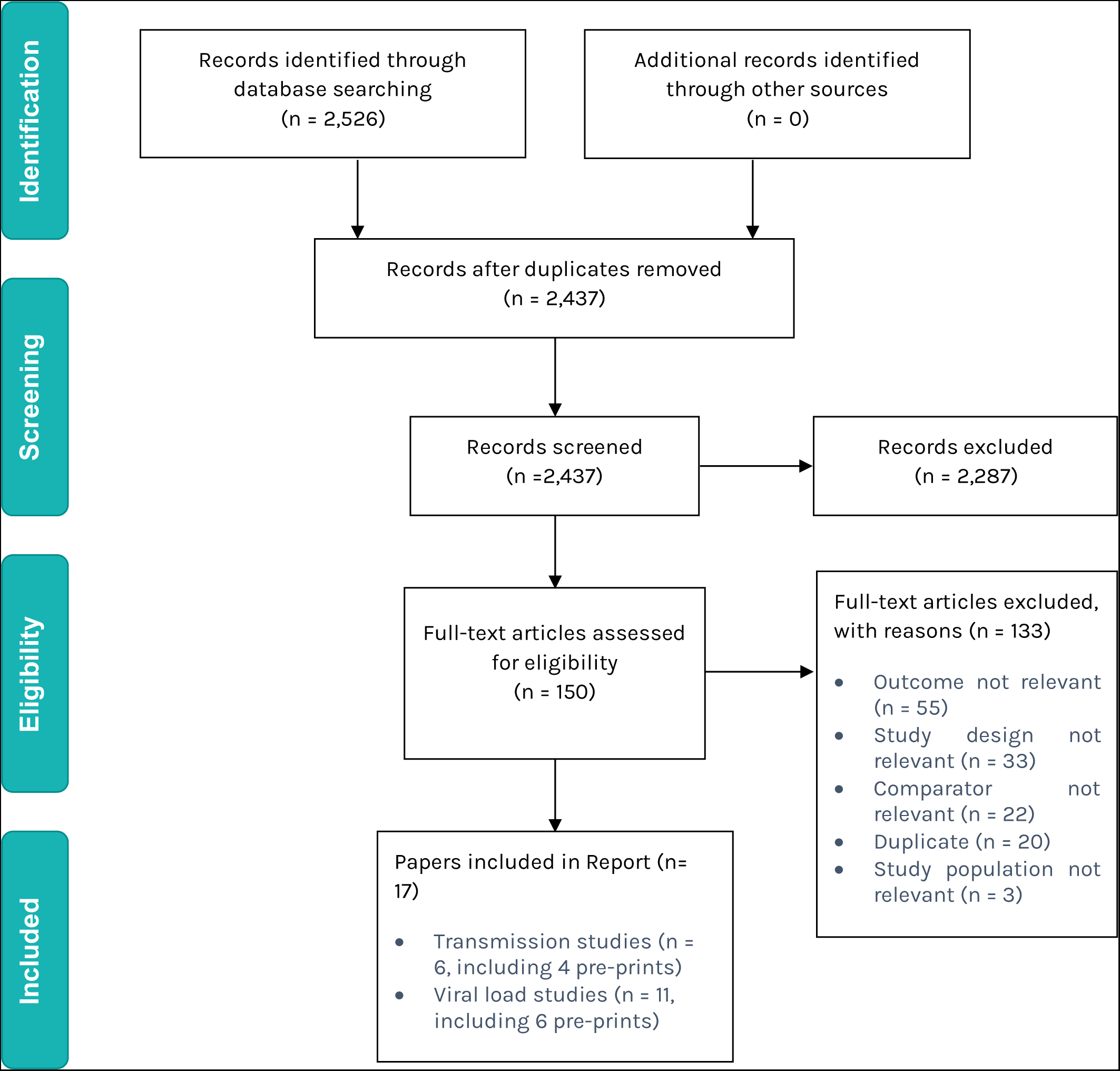
PRISMA diagram (for updated review)

### Data extraction and risk of bias assessment

Data extraction was completed by 1 reviewer and checked by a second. Only results directly relevant to the review questions were extracted.

Studies were assessed using the quality criteria checklist (QCC) for primary research (Academy of Nutrition and Dietetics 2016). This risk of bias tool can be applied to most study designs (observational and interventional) and is therefore suitable for rapid reviews of mixed type of evidence. It is composed of 10 validity questions based on the criteria and domains identified by the Agency for Healthcare

Research and Quality to assess the methodological quality of a study (that is, the extent to which a study has minimised selection, measurement and confounding biases) (West et al. 2002). In the QCC tool, 4 questions are considered critical (on selection bias, group comparability/confounding, interventions/exposure and outcome). A study will be rated as high quality if the answers to the 4 critical questions are ‘yes’ (and at least one additional ‘yes’). The study will be rated as low quality if 2 or more of the critical questions are answered ‘no’ and/or if ≥50% of the remaining questions are answered ‘no’. Otherwise, the study will be rated as medium quality. Judgments were made on case by case for questions answered as ‘unclear’. To note that we report these ratings as ‘quality’ ratings for consistency with the name of the tool, although here quality needs to be understood as ‘methodological quality’ as part of a risk of bias assessment.

QCC ratings are reported in the data extraction tables, **Supplementary Tables 1 and 2**.

Variations across populations and subgroups, for example cultural variations or differences between ethnic, social or vulnerable groups were considered, where evidence was available.

## 11. About the Wales COVID-19 Evidence Centre

The WCEC integrates with worldwide efforts to synthesise and mobilise knowledge from research. We operate with a core team as part of Health and Care Research Wales, are hosted in the [Wales Centre for Primary and Emergency Care Research (PRIME)](http://www.primecentre.wales/), and are led by [Professor Adrian Edwards of Cardiff University](https://www.cardiff.ac.uk/people/view/123022-edwards-adrian).

The **core** team of the centre works closely with collaborating partners in [Health Technology Wales](https://www.healthtechnology.wales/), Wales Centre for Evidence-Based Care, [Specialist Unit for Review Evidence centre](https://www.cardiff.ac.uk/specialist-unit-for-review-evidence), [SAIL Databank](https://healthandcareresearchwales.org/about-research-community/secure-anonymised-information-linkage-sail-databank), Bangor Institute for Health & Medical Research/ Health and Care Economics Cymru, and the [Public Health Wales Observatory](https://phw.nhs.wales/services-and-teams/observatory/).

Together we **aim** to provide around 50 reviews per year, answering the priority questions for policy and practice in Wales as we meet the demands of the pandemic and its impacts.

**Director:**

Professor Adrian Edwards

**Contact**

**Website including report library:** https://healthandcareresearchwales.org/about-research-community/wales-covid-19-evidence-centre

**Prepared by:** Jessica Williams, Lauren Elston, Jenni Washington and Thomas Winfield from [Health Technology Wales](https://healthtechnology.wales/)

